# Learning the electronic health record at the minute-scale

**DOI:** 10.64898/2026.10.04.26364562

**Authors:** Kush Narang, Newton Addo, Jaskaran Bains, Maytal Firnberg, Sonny Tat, Anneka Hooft, Daniela Chanci Arrubla, Baraa Abed, Chandan Singh, Rishikesan Kamaleswaran, Jean Feng, Aaron Kornblith

## Abstract

Electronic health record foundation models are traditionally trained on the scale of years, days, or hours. These timescales, however, lack the minute-scale resolution required to directly guide clinical decisions at the bedside. We introduced minute-scale learning, a new paradigm for training and evaluating models, and developed MINT, a minute-scale foundation model for pediatric emergencies. MINT was pretrained and validated on 766,733 pediatric emergency department visits at five health systems, comprising 16 years of data from 10 hospitals. On minute-scale forecasting tasks, MINT outperformed and generalized to external health systems better than task-specific models (superior in 21 of 25 comparisons). MINT outperformed physicians in forecasting escalations of respiratory support. MINT demonstrated uniquely minute-scale capabilities including department-scale monitoring, dynamic risk explanations, individualized physiologic response forecasts, and hypothesis generation. Learning at the minute-scale improves performance, strengthens generalizability, and provides insights, actionability, and scientific capabilities that are not accessible at other timescales.

## Introduction

Foundation models trained on electronic health records (EHRs) have advanced rapidly, but most learn at timescales that are too coarse to directly influence clinical decisions^1^. Most foundation models, predominantly trained on diagnosis codes, represent patients on the scale of months to years and forecast outcomes over a patient’s lifetime^2–9^. A smaller group of acute care foundation models function on the scale of hours to days, typically forecasting deterioration and interventions between a few hours and a week ahead^10–14^. Though these approaches are useful for epidemiological studies and coarse risk stratification, existing training and evaluation paradigms lack the minute-scale resolution needed to guide clinical decision-making at the bedside.

In acute care settings, clinicians rapidly evaluate new findings to dynamically update resuscitation strategies. Even slight delays in recognizing decompensation or initiating interventions are often associated with worse outcomes^15–17^. Without minute-scale models, clinicians rely on individual gestalt, monolithic guidelines, underpowered decision-support tools, or more recently, large language models (LLMs), all of which are susceptible to bias, difficult to adapt to local practice, and insufficiently precise for the sickest patients^18–27^. The development of higher-resolution foundation models may be transformative in emergency and critical care practice settings that carry significant morbidity and mortality burdens that contribute to up to half of all years of life lost^28–30^. We selected pediatric emergency care as our case study to introduce minute-scale learning, defined as training models on sequences that preserve minute-level information to forecast, explain, and manage near-term changes in health. Pediatric emergency care offers a sharp evaluation because children arrive undifferentiated, case severity varies widely, and age-dependent physiology introduces new complexity. Moreover, because decompensation is rare and practices vary across hospitals^31,32^, most sites lack sufficient data to train custom models, creating demand for decision support that generalizes. Most importantly, half of childhood deaths occur within hours of emergency department arrival, and a significant fraction are likely preventable with improved readiness^31,32^.

In this study, we introduce minute-scale learning and present MINT (MINute-scale Trajectory model), a foundation model pretrained and validated on 766,733 pediatric emergency department (ED) visits across ten hospitals spanning five health systems. MINT achieved state-of-the-art performance in forecasting minute-scale deterioration while generalizing to external sites more effectively than traditional models in an EHR transfer. We further evaluated the minute-scale paradigm with six experiments, inspired by previous literature, to demonstrate capabilities only learnable at the minute-scale: forecasting short-horizon interventions against physicians and a frontier LLM^33,34^; performing continuous monitoring across several patients^35–38;^ providing dynamic, individualized risk explanations^34^; learning clinically-sensible physiologic associations^39^; estimating individual responses to interventions^6,40–42;^ and, replicating the results of previous registry-scale studies^43,44^. Our findings demonstrate the unique capabilities of minute-scale learning and introduce new frontiers for clinical actionability, operational utility, and scientific inquiry with EHR models.

## Results

To learn clinical trajectories at the minute-scale, we developed MINT, an autoregressive, decoder-only transformer based on Delphi-2M^5^, an adapted GPT-2 architecture (**Figure 1a**). MINT was trained on 283,682 pediatric ED visits from 2011 to 2023 at three University of California, San Francisco (UCSF) hospitals, evaluated internally on 81,285 temporally held-out visits from 2024 to 2025 at UCSF hospitals, and externally validated on 401,766 visits at seven hospitals across four other University of California (UC) health systems (**Figure 1c**). Visit characteristics are available in **Extended Data Table 1**.

**Figure 1.**
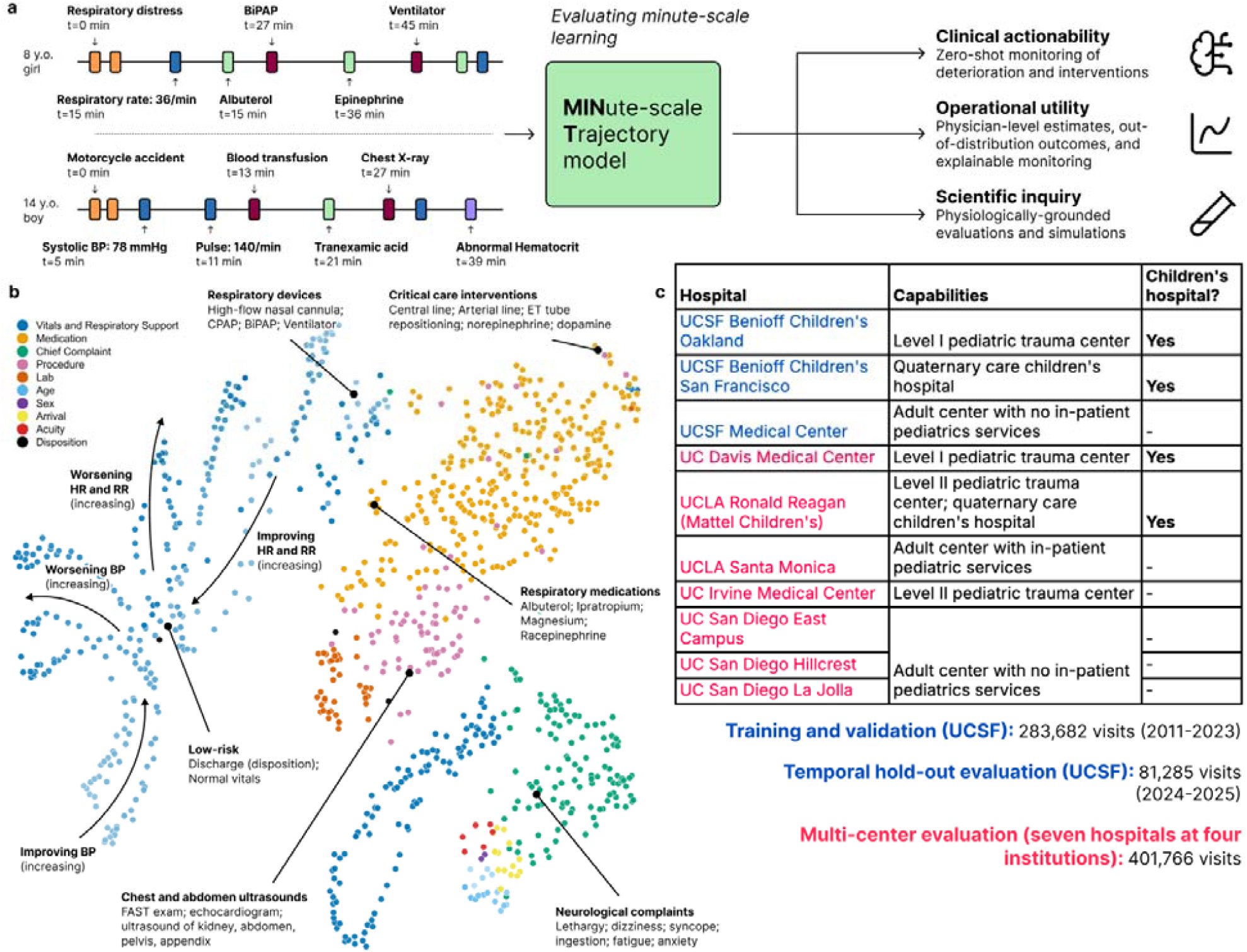
A minute-scale foundation model. Overview of the MINT model, its learned representations, and the multicenter study design. **Panel A**, mock pediatric emergency department trajectories illustrating heterogeneous longitudinal sequences of vital signs, medications, laboratory results, imaging, procedures, and clinical events that are integrated into a unified patient representation to enable minute-scale inference for treatment and physiologic forecasting. These clinically grounded representations support applications in real-time monitoring, operational decision-making, and model interpretability. **Panel B**, two-dimensional visualization (UMAP) of learned token embeddings for common tokens (300+ occurrences), demonstrating that MINT organizes diverse clinical concepts into semantically meaningful clusters, with related token types occupying nearby regions of the representation space. **Panel C**, overview of the study cohorts and participating health systems. Children’s hospitals are defined as those with membership in the Children’s Hospital Association. Numbers indicate the total visits included in the training, temporal hold-out, and multicenter evaluation cohorts.

### Forecasting at the minute-scale

We first evaluated whether MINT could forecast five physiologic deterioration outcomes, adapted from age-specific Pediatric Advanced Life Support (PALS) guidelines, and five intervention outcomes based on the airway, breathing, and circulation domains of pediatric resuscitation (**Extended Data Table 2**).

Across these minute-scale tasks, zero-shot MINT achieved a mean AUROC of 0.93 (95% CI, 0.92-0.93) and AUPRC of 0.36 (95% CI, 0.34-0.39) despite low incidence (median [IQR] incidence, 1.6% [0.5%-3.8%]) (**Figure 2a**, **Supplementary Figure 1**). MINT exceeded both task-specific models for eight of 10 outcomes and was equivalent for one; it was inferior only to XGBoost for airway rescue medications, the highest-incidence intervention outcome (**Figure 2**).

**Figure 2.**
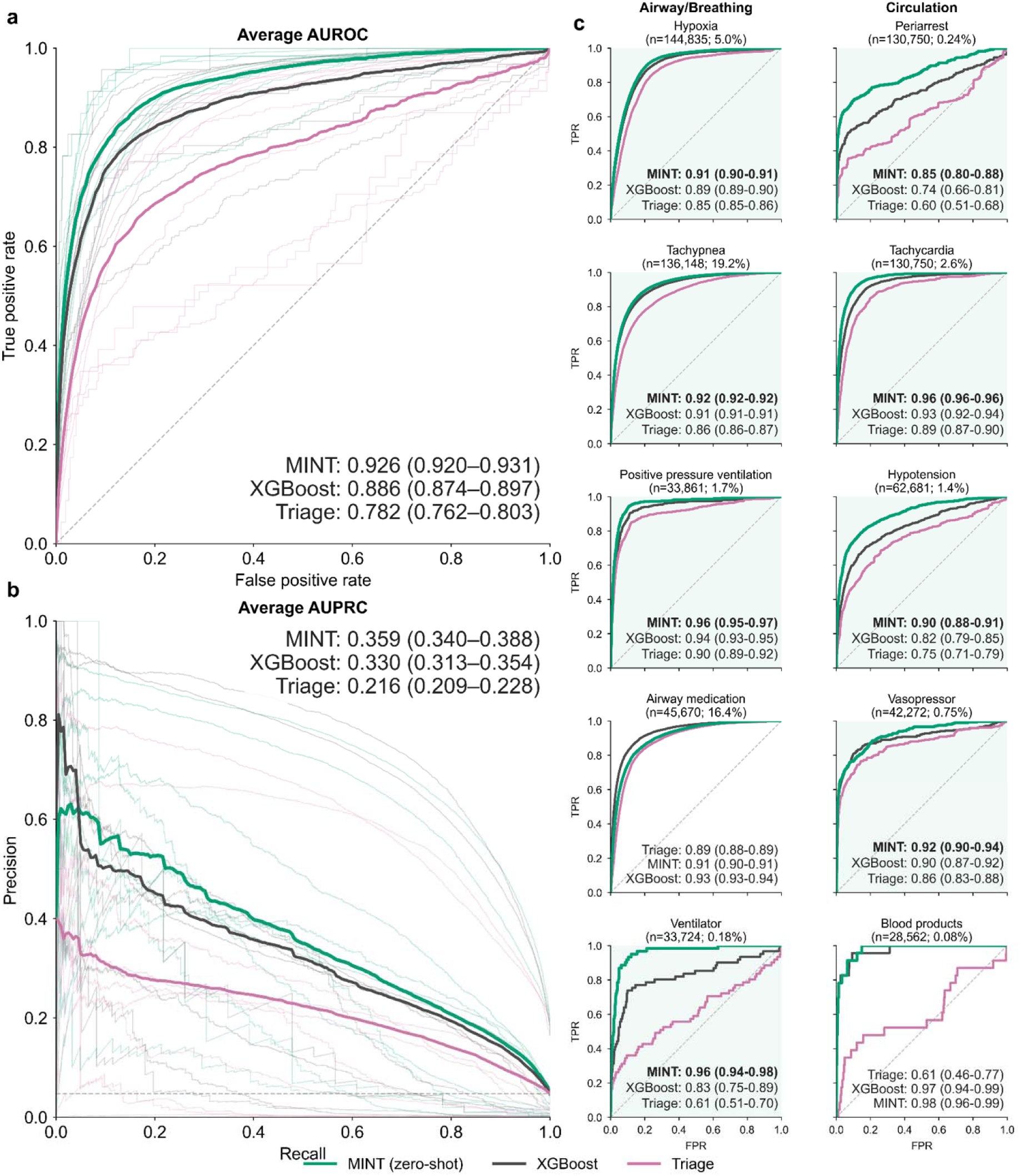
Minute-scale forecasting with MINT. MINT performance across 10 physiologic deterioration and intervention outcomes compared against XGBoost (black) and triage (pink) task-specific models. **Panel A and B**, receiver operating characteristic (ROC) curves (**Panel A**) and precision-recall curves (**Panel B**), macro-averaged across outcomes. **Panel C**, ROC curves for individual airway/breathing (left column) and circulation outcomes (right column). Numbers in each mini-panel title indicate the evaluation cohort size and outcome incidence. Values are shown with 95% confidence intervals (CIs). The light green background denotes a significant improvement delivered by MINT over other methods, defined as 95% CIs for the paired AUROC difference excluding zero.

MINT maintained discriminative performance when outcome thresholds were redefined without re-training (**Supplementary Figure 2**). To evaluate data efficiency in learning, we trained a linear probe on MINT and found that it matched XGBoost for forecasting airway rescue medications with between 5-fold and 10-fold fewer training cases at a high AUROC (**Supplementary Figure 3**). MINT’s performance was consistent across demographic groups, though varied by age group for two outcomes (**Supplementary Figure 4**).

To understand which design choices drove performance gains, we progressively ablated tokenization timescale (minute-scale, hour-scale, and day-scale) and token types. These ablations identified minute-scale tokenization and vital signs as the largest contributors of superior performance (**Extended Data Figure 1**). Switching timescales from just minutes to hours resulted in significant performance decreases for six of the 10 outcomes.

### Generalizing to external health systems

A frequently cited benefit of foundation models is their ability to generalize across institutions better than traditional methods, though this often goes untested in existing literature^45–47^. To address this gap, we evaluated MINT’s ability to generalize to four external institutions, grouped into five analytic sites; these sites represent seven UC hospitals that span a wide range of pediatric capabilities (**Figure 1c**).

Our external generalizability setup involved a full re-tokenization process (e.g., token remapping) from an Epic-based to an OMOP-based EHR schema (**Methods** and **Supplementary Tables 1-2**). This process left more than one-third of tokens unmapped, simulating the friction and potential vocabulary loss during a real-world model transfer. We compared MINT (trained on UCSF cases) against site-agnostic (trained on UCSF cases) and site-specific (trained separately on each site’s cases) XGBoost comparators.

Across the five external sites, zero-shot MINT exceeded site-agnostic models in 21 of 25 site-outcome comparisons and was equivalent in the remaining four, without site-specific or task-specific training (**Figure 3a**, **blue**). Against site-specific models, MINT matched or exceeded local performance in all 10 comparisons at the two lower-volume sites (<20,000 visits) but was inferior in seven of 15 comparisons at the three higher-volume sites (>50,000 visits) (**Figure 3a**, **pink**). MINT outperformed site-specific models for peri-arrest at five (100%) and hypotension at four (80%) of five sites, the two most severe physiologic outcomes. MINT’s advantage over site-specific models was inversely associated with the number of positive training cases (ρ=-0.91; **Figure 3b**, **Supplementary Figure 5**).

**Figure 3.**
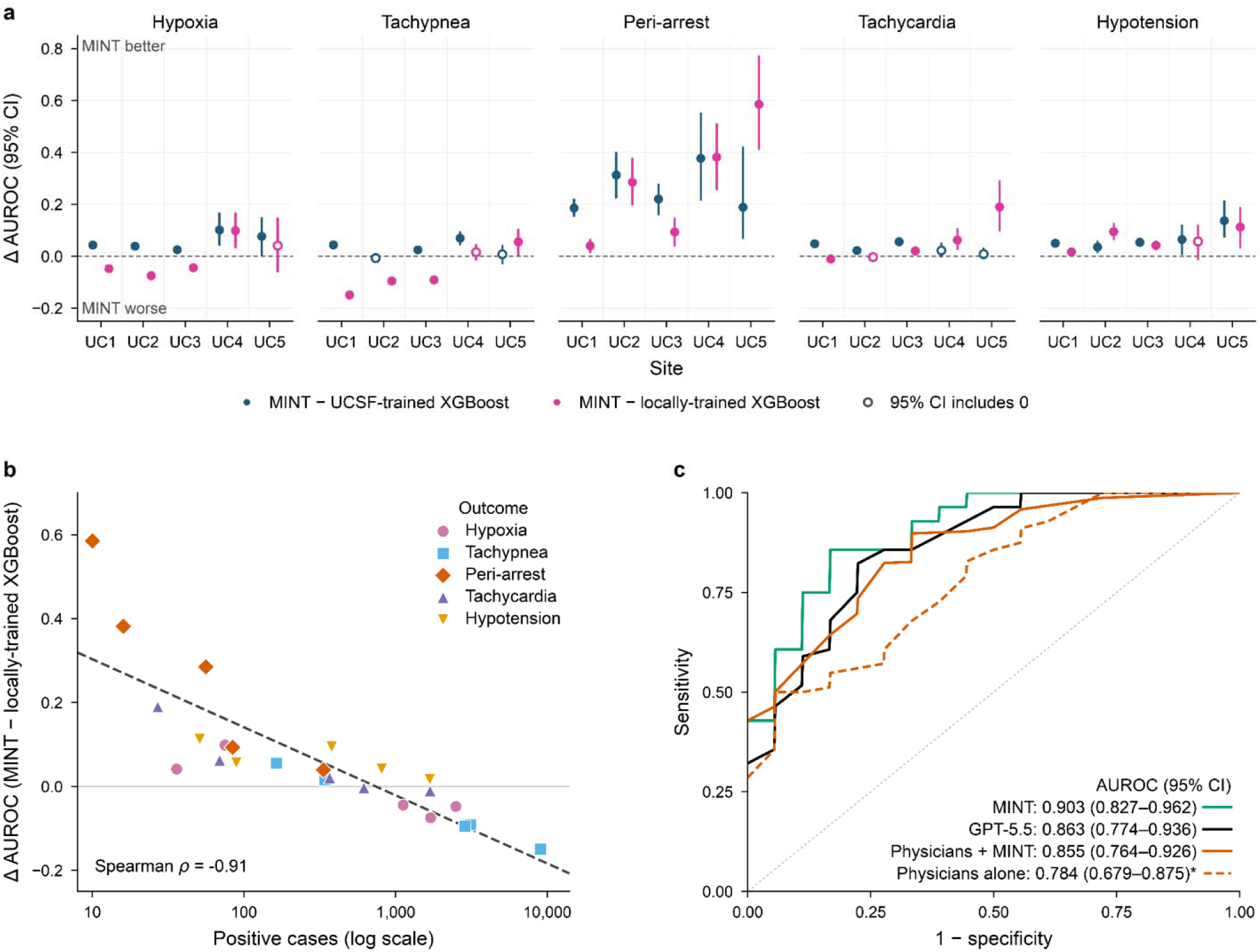
Generalization to external health systems and forecasting performance against physicians and an LLM. AUROC improvements of zero-shot MINT over XGBoost models across external sites, representing four health systems and seven hospitals. **Panel A**, comparisons between MINT and site-agnostic XGBoost models (blue) and site-specific XGBoost models (pink). **Panel B**, performance differences between MINT and site-specific XGBoost models versus the number of positive outcome samples with a reported Spearman rank correlation. **Panel C**, AUROC curves for MINT, GPT-5.5 (in-context learning), and four board-certified emergency medicine physicians (with MINT and without) for forecasting positive pressure ventilation initiation in the next 5 to 60 minutes. *denotes a significant performance difference compared to MINT alone, defined as 95% CIs for the paired AUROC difference excluding zero.

We next considered whether site-specific training (e.g., continued pre-training, fine-tuning, or transfer learning) could further improve MINT’s performance. Training site-specific linear probes on MINT reduced the number of inferior comparisons from seven to three of 25, all for tachypnea (**Extended Data Figure 2a**). Zero-shot MINT, linear probes on MINT, and linear probes on fine-tuned MINT (e.g. after continued pre-training) generally delivered the most improvements over XGBoost comparators (**Extended Data Figure 2a**). The most complex design, fine-tuned end-to-end MINT (e.g., continued pre-training with task-specific fine-tuning), delivered mixed performance results spanning some gains at high-volume sites to massive losses at low-volume sites (**Extended Data Figure 2c**). For task-specific methods, performance was positively correlated with the number of positive cases available for site-specific training (Pearson r, +0.81 for end-to-end fine-tuning and +0.66 for site-specific XGBoost) (**Extended Data Figure 2b**). Additional data from these experiments are available in **Supplementary Figure 5**.

### Forecasting positive pressure ventilation versus physicians and LLMs

Beyond forecasting performance, we next considered whether MINT could both perform and assist clinical decision-making, an ability that is often implied but untested in EHR foundation models^1,48^. We compared MINT’s ability to forecast the initiation of positive pressure ventilation (PPV) within the next 60 minutes against board-certified physicians and GPT-5.5^49^. PPV was selected as the outcome because it is uniquely monitored at the minute-scale and early initiation is associated with improved outcomes^50–52^.

In a subset of children with severe respiratory distress (PPV incidence, 28%), MINT achieved an AUROC of 0.90 (95% CI 0.83-0.96), exceeding physicians alone (0.78, 95% CI 0.68-0.88; AUROC difference, -0.12; 95% CI, -0.21 to -0.04) (**Figure 3c**). Physicians with MINT were more accurate than physicians alone, though not significantly (0.86; 95% CI 0.76-0.93; AUROC difference 0.07; 95% CI -0.01 to 0.16).

MINT performed comparably to GPT-5.5 (0.86; 95% CI 0.77-0.94) while running approximately 800 times faster and orders of magnitude lower cost per case (**Extended Data Table 3**); GPT-5.5 took an average of 8.3 seconds and cost 3.7 cents per predictive case (**Extended Data Table 3**). For an ED that sees 100 children a day and seeks to run risk estimates every few minutes (e.g., 100 times per child), deploying GPT-5.5, for this single task, would cost an estimated $370 per day, totaling 10,000 inference runs. We also tested GPT-5-mini^53^ which cost 0.22 cents per case, which would extrapolate to $22 a day in the hypothetical ED deployment. While MINT did not significantly outperform GPT-5.5, it did significantly outperform GPT-5-mini (AUROC difference, -0.12; 95% CI -0.21 to -0.04).

### Generalizing to out-of-vocabulary outcomes

Another important benchmark for foundation models is the ability to generalize to new outcomes in data-limited regimes^10,46,47^. We benchmarked MINT on this axis by training linear probes for hospital admission, intensive care unit (ICU) admission, sepsis, and septic shock.

MINT linear probes significantly outperformed XGBoost for three outcomes and performed comparably on forecasting admission (**Extended Data Figure 3**). We also tested ETHOS, an adult EHR foundation model, for forecasting hospital admission using the first four hours of ED data and found that it could not perform better than random chance (**Supplementary Table 3**).

Comparing MINT to XGBoost, the largest performance gains were for septic shock (AUROC MINT probe 0.88 [0.84-0.91] vs XGBoost 0.74 [0.69-0.80]) (**Extended Data Figure 3a**). At 90% sensitivity (an established clinical operating threshold), MINT achieved a number needed to evaluate (NNE, [1 / positive predictive value]) of 22 for sepsis and 55 for septic shock, compared to XGBoost’s NNE of 55 and 91, respectively. MINT’s performance was comparable to a state-of-the-art sepsis forecasting algorithm^54^, despite MINT’s vocabulary and pre-training containing no information on diagnosis codes or chronic illness. These findings suggest that MINT may provide data efficiency and performance benefits for rare event outcomes.

### Monitoring a virtual emergency department continuously

We next considered whether MINT’s training paradigm enabled novel capabilities only learnable at the minute-scale. We first evaluated continuous monitoring of many patients simultaneously, an important capability in emergency medicine that has been tested in previous machine learning studies^34^ but not frequently in foundation models.

In a virtual ED simulation, MINT identified the children most likely to be admitted within minutes of arrival and refined rankings as trajectories accrued (**Figure 4a-b**). Across 1,000 simulations, children ranked highest at 30 minutes had higher admission rates, longer lengths of stay, and greater medication, procedure, and laboratory use (**Figure 4c**). Low-ranked children who were nonetheless admitted either decompensated later in the ED visit or had chronic comorbidities not represented in the MINT vocabulary.

**Figure 4.**
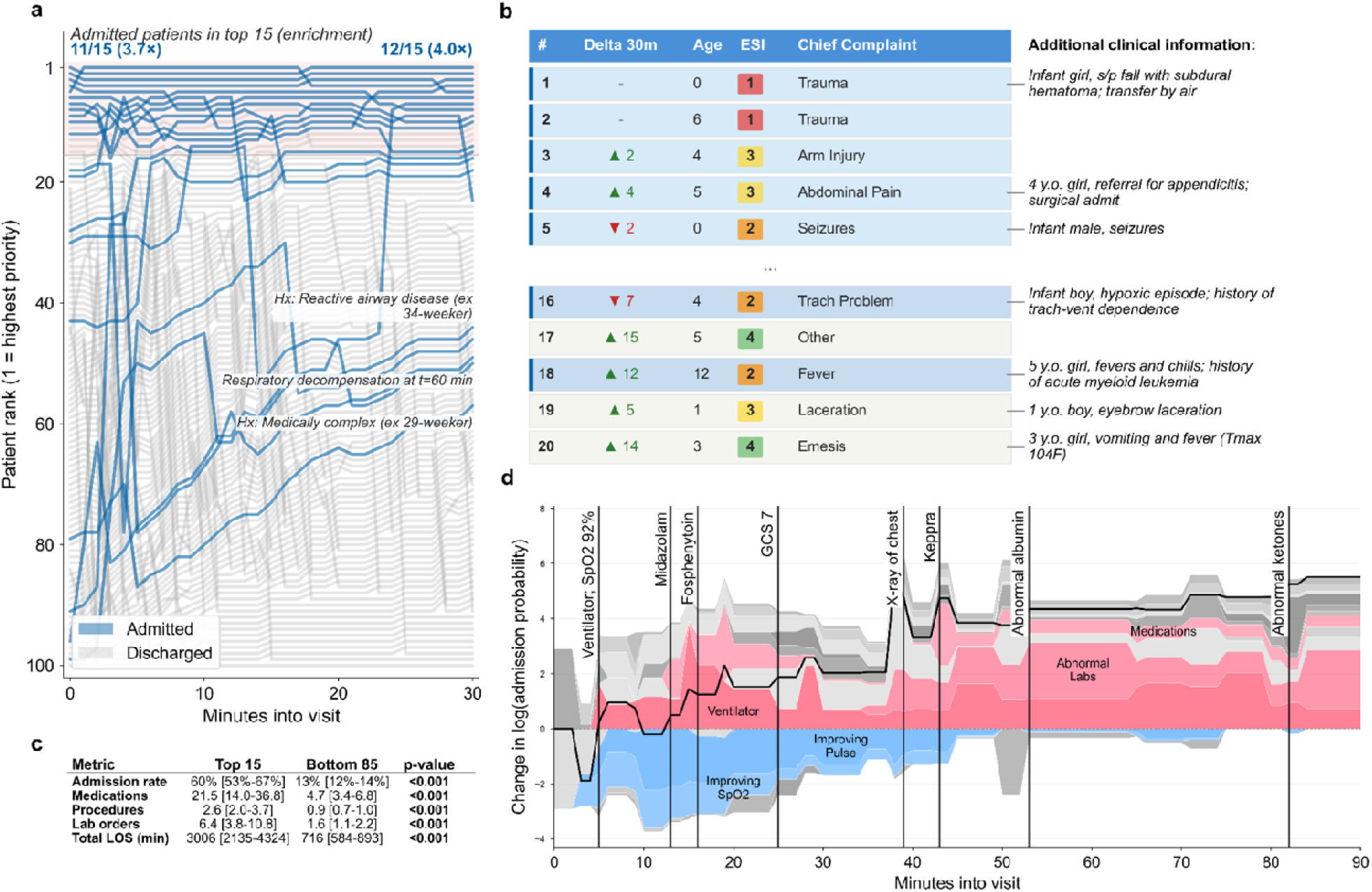
Continuous minute-scale monitoring and dynamic risk explanations with MINT. Applications of MINT for real-time operational monitoring and individualized risk assessment. **Panel A**, simulation of a pediatric emergency department with 100 children continuously reprioritized during the first 30 minutes of the emergency department (ED) visit, ranked by MINT’s probability of admission. Blue trajectories denote children ultimately admitted, and the shaded region indicates the highest-priority 15 children. Representative trajectories illustrate dynamic reprioritization as new clinical information becomes available. **Panel B**, an example ED track board after 30 minutes showing changes in priority together with demographic information and additional details. **Panel C**, results from 1,000 simulated EDs comparing resource utilization between the top 15 ranked children versus the bottom 85 at 30 minutes. Values are presented as median [IQR] across simulations with paired Wilcoxon signed-rank tests on simulation-level differences. **Panel D**, dynamic explanation of admission risk for an 8-year-old girl with seizures and respiratory failure, estimated with the cumulative density function (12-hour time horizon). The black line shows the estimated change in log probability of admission over time, and the stacked colored regions represent cumulative contributions of token groups. Vertical lines indicate key clinical events, demonstrating how medications, procedures, physiologic abnormalities, and laboratory findings dynamically alter predicted risk.

### Generating context-dependent risk explanations

We next tested whether MINT could generate dynamic risk explanations, a well-established and clinically important capability of past machine learning methods^34^.

MINT generated minute-by-minute risk explanations for a critically ill eight-year-old girl with seizures and respiratory failure (**Figure 4d**). Additional cases are shown in **Extended Data Figure 4**. Medications, procedures, and abnormal laboratory results were generally associated with the largest jumps in risk, while vital signs had more modest effects. Advanced respiratory support was associated with increased risk, whereas blow-by oxygen and room air tokens were associated with decreased risk, suggesting that MINT learned the intensity ladder of respiratory support.

The contribution of an individual token depended strongly on its clinical context rather than its identity alone. For example, midazolam substantially increased risk for the 8-year-old girl presenting with seizures (**Figure 4d**), consistent with treatment of a critically ill patient, but produced little change in the boy who received it as procedural sedation for laceration repair (**Extended Data Figure 4a**). Likewise, acquisition of an electrocardiogram (ECG) increased risk during evaluation of an infant girl with respiratory distress but had little effect in the adolescent with SVT, where cardiac evaluation was practically a clinical certainty (**Extended Data Figure 4b-c**). These cases suggest that MINT may be able to use context to learn the clinical indication for medications and procedures rather than assigning uniform risk.

### Evaluating physiologic associations and precision capabilities

We finally considered whether MINT could learn clinically sensible physiologic associations, estimate individual responses to interventions, and replicate the results of registry-scale studies, three important directions established by prior work that we tested^6,39,40,43,44^. Of note, MINT is not a causal model, and none of our results are intended to imply or demonstrate causation. Instead, we were broadly interested in whether minute-scale foundation models are capable of initial progress towards these important directions.

Under simulated counterfactual interventions, MINT reproduced expected vital-sign associations for single-token interventions (pulse after epinephrine, +4.6 bpm for one dose and +7.4 bpm for three; oxygen saturation after respiratory support, +0.9 percentage points) and for composite, multi-token interventions (Glasgow Coma Scale score after rapid sequence intubation, -3.3 points; systolic blood pressure [SBP] after massive transfusion, +5.8 mmHg) (**Figure 5a**). MINT also inverted several associations, forecasting a decreased respiratory rate after naloxone, decreased SBP after dopamine, and increased pulse after adenosine; in these cases, it reproduced the indication rather than the therapeutic effect, a pattern of confounding by indication (**Supplementary Figure 6-7**). MINT more accurately recovered the expected direction of effect for medications that were more frequently represented in the ED training data (odds ratio, 7.0 [95% CI, 1.1–44.2] for every 10-fold increase in the number of training examples).

**Figure 5.**
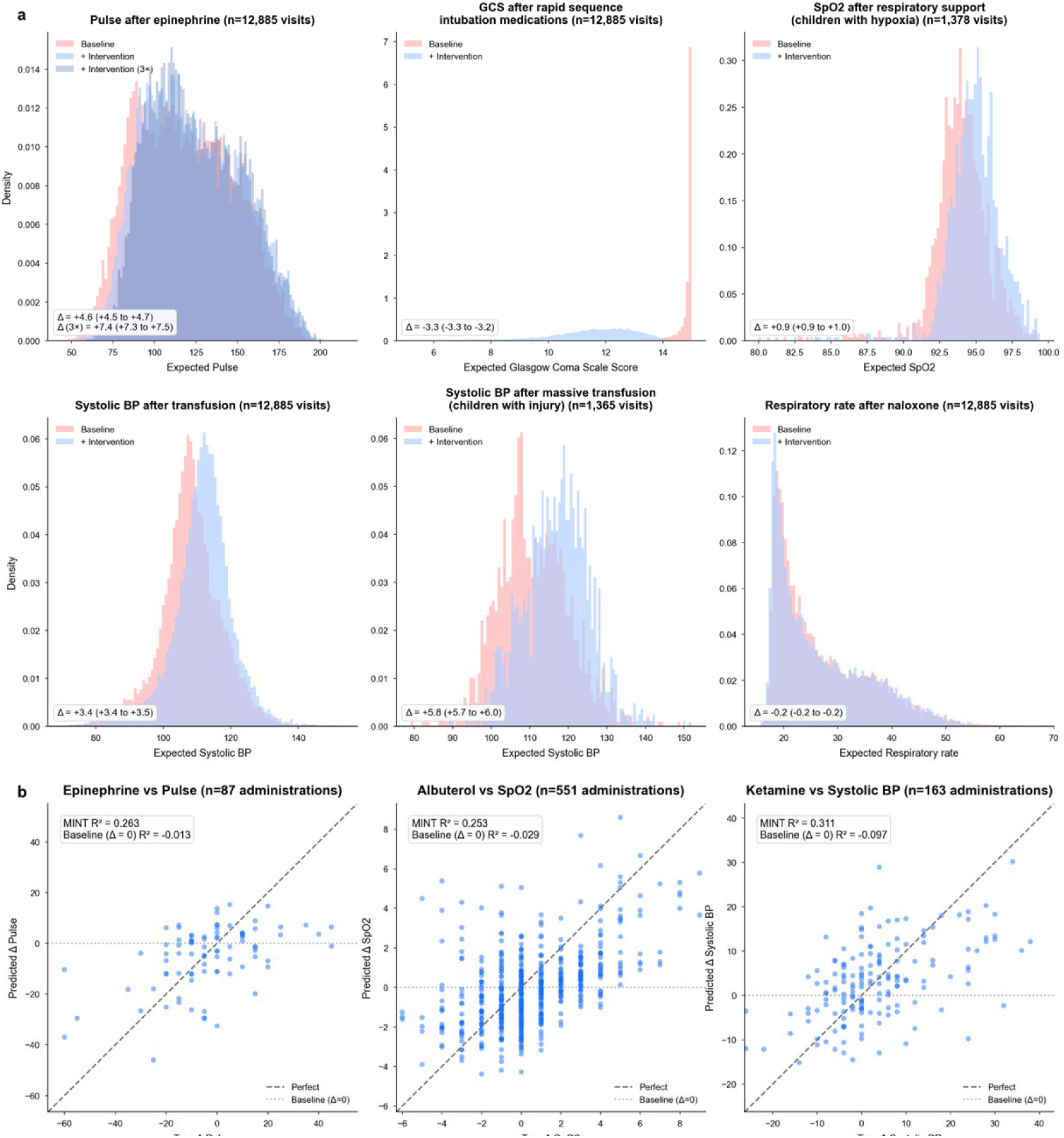
Physiologic interpretability and estimating individual responses with MINT. MINT learned clinically meaningful associations between interventions and physiology and leveraged these representations to forecast individualized physiologic responses. **Panel A**, distributional shifts in predicted physiologic states after synthetically adding or removing clinically relevant intervention tokens to patient trajectories. Histograms compare vital signs distributions before (red) and after intervention (blue). Mean predicted change (Δ) is reported with bootstrapped 95% confidence intervals and p-values calculated with Wilcoxon signed-rank tests. **Panel B**, predicted versus observed physiologic responses among children who received interventions of interest in the emergency department with before and after vital signs. The baseline (naive) model predicted the most recent vital sign before the intervention. For both analyses, vitals were forecasted with a weighted softmax over the next-token logits for the vital sign type of interest (e.g., forecasting pulse by taking the weighted probability over all next-token predicted pulse tokens).

In individual children who received an intervention, MINT forecast the observed vital-sign change with modest accuracy (pulse after epinephrine, R^2^=0.26), oxygen saturation after albuterol (R^2^=0.25), and SBP after ketamine (R^2^=0.31), capturing changes in both directions rather than always predicting the canonical drug effect (**Figure 5b**).

MINT token embeddings also organized into clinically coherent clusters, with nearest neighbors and vector analogies recovering relationships across token types (**Figure 1b**; **Extended Data Figure 5**). MINT produced clinically coherent analogies across multiple token categories (**Extended Data Figure 5b**). MINT learned that akin to the relationship between orthopedic reductions and ketamine, intubation was related to propofol, rocuronium, and lorazepam. In both cases, these medications are typically delivered before the intervention of interest. Midazolam, epinephrine, and lorazepam were identified as seizure treatments, akin to acetaminophen as a fever treatment. Febrile seizure was associated with younger children, while (non-febrile) seizure was associated with older children. Transfusion and tranexamic acid, two critical hemorrhage control interventions, were associated with airway obstruction and injury complaints.

MINT also learned gradient structures for continuous vital signs with manifolds of increasing values passing through the low-risk centroid at the same location as the “discharge” token (**Figure 1b**). These findings suggest that MINT learned clinically meaningful concepts from the self-supervised pre-training objective.

### Generating hypotheses with counterfactual simulations

Finally, we examined whether counterfactual simulations with MINT could recover the treatment-outcome associations from two recent registry-scale studies reporting benefits from earlier and more aggressive ED interventions. Though MINT is not capable of providing true causal estimates, we hypothesized it could demonstrate early capabilities for hypothesis generation. We considered the established association between decreased hospitalization rates in acute gastroenteritis treated with ondansetron (Ramgopal et al. 2026; n=543,541) and sickle cell vaso-occlusive crisis treated with early opioid analgesia (Gwarzo et al. 2025; n=2,538).^55,56^

After replicating the original inclusion criteria, we counterfactually removed tokens from each trajectory and determined the proportion of children with increased probability of admission (**Table 1**); increased admission probabilities without the intervention would suggest that the intervention was protective. Counterfactual removal of ondansetron in gastroenteritis cases (n=1,000) increased predicted admission probability in 91.7% of children (**Table 1**). Removal of early analgesia in vaso-occlusive crisis (n=53) similarly increased predicted admission probability in 94.3% of children (**Table 1**). These simulations reflected lower risk with more aggressive intervention and matched the direction of both original studies, though we did not estimate or compare effect sizes.

**Table 1.** Counterfactual simulations for hypothesis generation with MINT. Published associations with hospitalization are compared with MINT predictions after counterfactual removal of clinical interventions. Ondansetron was associated with lower odds of hospitalization in a registry-scale analysis of 543,541 children (Ramgopal et al. 2026; odds ratio [OR] 0.61)^56^. Counterfactual removal of ondansetron increased predicted admission probability in 91.7% of 1,000 children with acute gastroenteritis. Early opioid administration was associated with lower odds of hospitalization in a registry-scale analysis of 2,538 children (Gwarzo et al. 2025; OR, 0.84).^55^ Counterfactual removal of early pain management increased predicted admission probability in 94.3% of 53 children with sickle cell crisis. Bars indicate the proportion of children with an increase in predicted admission probability, rather than the magnitude of that increase. These simulations demonstrate directional agreement with published associations and support hypothesis generation; they do not establish causal treatment effects.

| Intervention | Population | Published study |  |  | MINT simulation,<br>removing intervention |  | Directional<br>agreement |
| --- | --- | --- | --- | --- | --- | --- | --- |
|  |  | Reference | Study size<br>N | Hospitalization<br>OR | n | Increased predicted<br>admission risk<br>(% of visits) |  |
| Ondansetron | Children with acute<br>gastroenteritis | Ramgopal et al.<br>(2026) | 543,541 | 0.61 | 1,000 | 91.7% | ✓ |
| Early pain<br>management | Children with sickle<br>cell pain crisis | Gwarzo et al.<br>(2025) | 2,538 | 0.84 | 53 | 94.3% | ✓ |

Both experiments were chosen because the correct answer lies in the opposite direction of confounding by indication; children receiving ondansetron or early analgesia are more symptomatic at presentation. A possible explanation for why confounding is less pronounced here is because ondansetron and analgesia are regularly administered to children of all severity levels, while more confounded interventions like dopamine and adenosine are exclusively used in profoundly ill children (**Figure 5**; **Supplementary Figure 6**). Another potential explanation is that these interventions are administered more frequently and learned as more general interventions, consistent with findings across 26 medication-outcome pairs based on the *American Academy of Pediatrics* care guidelines^57^ (**Supplementary Figure 7**).

## Discussion

We introduced minute-scale learning and developed MINT, a minute-scale foundation model that learns from trajectories of pediatric emergencies. MINT outperformed traditional methods for critical forecasting tasks, strongly generalized to four external health systems in a simulated EHR transfer, and extended to out-of-vocabulary, rare event outcomes. MINT demonstrated novel capabilities for a foundation model, including monitoring across the department, explaining individual risk, forecasting responses to interventions, and matching the directional findings of registry-scale studies. These findings establish minute-scale learning as a viable paradigm and carry important implications for clinical teams and machine learning researchers alike.

The key technical advance in MINT is learning clinical trajectories at the minute-scale, the resolution at which clinicians manage resuscitation and make emergency care decisions. With this new approach, MINT transitions foundation models from broad, population-level prognostic tools^1,5,8^ into individualized, clinically actionable bedside assets. MINT outperformed physicians for forecasting PPV initiation, an intervention associated with improved outcomes in children^50–52^. Additionally, physicians using MINT for decision support were more accurate than those without MINT, though this difference was not significant. To our knowledge, this is among the earliest evidence of a foundation model both surpassing and improving physician recognition of critical events on the minute-scale.

Forecasting individual physiologic responses to interventions could improve resuscitation outcomes^28,41,42,58–60^, while dynamic risk explanations could direct clinicians towards the reversible causes of deterioration^34,61,62^. Furthermore, by providing continuous monitoring across the ED, foundation models may act as real-time prioritization systems that could reduce mistriage-induced delays in care^63–66^.

The ED is a demanding setting for foundation models, and reasoning about life-saving clinical decisions at the minute-scale required extra physiology-grounded evaluations. Children arrive undifferentiated often without prior histories, documentation is sparse, and the outcomes that matter are rare, making deterioration a question of which child rather than when. Accordingly, we designed our minute-scale evaluations around experiments from prior literature to address the heterogeneous evaluation practices of current foundation model studies^45,46,48,67^. Though MINT generally learned correct treatment associations, our experiments revealed confounding by indication^68,69^, consistent with past findings that associative models are prone to learning spurious correlations and data shortcuts^70–73^. Notably, despite some confounding by indication, MINT successfully reproduced the directional associations identified in recent registry-scale studies of ondansetron and analgesia. These results suggest that, especially through the future integration of causal methods, foundation models may become valuable tools for hypothesis generation and *in silico* trials.

We further strengthened our evaluation with a robust multicenter evaluation that re-tokenized data from a new data model, leaving one-third of the vocabulary unmapped and reflecting the practical friction of transferring models across health systems^14,74,75^. This design addresses a recognized gap in multicenter validation studies of foundation models^46,48^. Despite vocabulary loss, MINT generalized better than task-specific models and outperformed site-specific models at smaller sites. MINT also matched state-of-the-art, purpose-built sepsis models on an out-of-distribution task, despite having a vocabulary that contained no diagnosis codes^54^. These results suggest that large clinical trial networks may benefit from investing in unified EHR datasets and developing foundation models that generalize across outcomes rather than focusing efforts on individual, task-specific methods.

MINT demonstrated strong performance across the spectrum of hospital capabilities, spanning large catchment, level I pediatric trauma centers to low-volume sites without inpatient pediatric services. Importantly, unlike existing billion-parameter LLMs and foundation models that require specialized hardware or corporate contracts to run securely^46,76,77^, MINT supported sub-second inference on consumer-grade laptops and fine-tuned on hundreds of thousands of visits in under 12 hours on a server with 16 GB of memory without a GPU, enabling deployment on the devices that community sites already have. This accessibility is important because it directly addresses the greatest paradox in clinical AI deployment: the sites with the greatest access to computational infrastructure are precisely the large academic centers that require the least amount of help to deliver high-quality care^46,76^.

This study has several limitations. First, all health systems studied are affiliated with the University of California. However, all five systems operate with distinct clinical practice guidelines, care for different geographic populations, and have varying levels of pediatric capabilities and expertise. We mitigated data similarity biases by re-tokenizing external data from an Epic-based to an OMOP-based data model to replicate the true complexity of multicenter transfer. Second, multicenter validation was restricted to only physiologic deterioration outcomes because clinical interventions were incompletely captured at external sites. Future studies will need to evaluate the generalizability of intervention forecasts. Third, due to date de-identification systems, small temporal overlaps between training and internal evaluation periods may exist for UCSF visits. Fourth, due to data privacy constraints, age was tokenized as years rather than months, which may obscure the steep physiologic changes in infancy. Fifth, physician comparisons were retrospective in a small cohort, and department-wide monitoring analyses were simulations. Prospective, randomized studies are necessary before making any conclusions on true clinical utility. Sixth, MINT is a purely associative model, making it unable to estimate causal effects and prone to confounding by indication. Future work should incorporate causal machine learning techniques into EHR foundation models to enable novel insights that are not possible with associative learning.

In this multicenter study, learning the electronic health record at the minute-scale improved forecasting and generalizability while enabling dynamic risk assessment, individualized physiologic predictions, and hypothesis-generating simulations in pediatric emergency care.

## Methods

We summarize the approach to training and evaluating MINT here; additional details are available in the **Supplementary Methods**. This study used fully de-identified data from the UCSF Clinical Data Warehouse (CDW) and the UC Health Data Warehouse (UCHDW) and did not require UCSF Institutional Review Board (IRB) review or informed consent. This study followed the TRIPOD+AI reporting guidelines^78^.

### Model architecture

MINT is based on the Delphi-2M architecture, with only limited changes to vocabulary size, block size, and most importantly, the token timescale, which we set to minutes since ED arrival rather than days since birth.

The final model was trained on a single NVIDIA L4 GPU, with additional inference performed on Apple silicon and CPU compute. All evaluations were zero-shot with no task-specific training unless otherwise stated and used softmax over the next-token distribution as the estimator, except for the feature importance analyses, which used exponentials to estimate event rates (**Supplementary Methods**). This contrasts with many previous methods that have relied on multiple trajectory rollouts for inference^1,11^.

### Development cohort and tokenization

MINT was trained using pediatric ED visits from 2011 to 2023 at three academic centers at the University of California, San Francisco (UCSF): two children’s hospitals (Benioff Children’s Hospital Mission Bay and Oakland), and one adult quaternary care hospital (UCSF Medical Center at Parnassus), accessed through the CDW. Visits were eligible if the child was younger than 18 years, and the ED disposition was “admit” or “discharge”; visits missing an ED physician note or with fewer than 12 tokens after processing were excluded.

All clinical data were processed into discrete tokens capturing the core elements of emergency care and resuscitation: triage details (age/sex, arrival method, acuity level), physiologic monitoring (vital signs), diagnostics and interventions (respiratory support, medications, procedures, abnormal laboratory results), and disposition (**Figure 1a**). Continuous values were rounded into bins for discrete tokenization; each token was linked to the minutes since ED arrival at which the event occurred, and tokens occurring fewer than 25 times across the dataset were removed.

### Generating forecasts for minute-scale outcomes

We evaluated 10 minute-scale forecasting tasks, five predicting physiologic deterioration and five predicting clinical interventions, based on the ABC domains of pediatric resuscitation. Physiologic outcomes were: hypoxia, tachypnea, peri-arrest (bradycardia with cardiopulmonary compromise), tachycardia, and hypotension, adapted from Pediatric Advanced Life Support (PALS) age-specific protocols. Intervention outcomes were: invasive mechanical ventilation, any positive pressure ventilation (PPV), airway rescue medications, vasopressors, and blood product transfusion. Full definitions for all outcomes are available in **Extended Data Table 2**.

For physiologic outcomes, we constructed a prediction case at every vital sign measurement, labeled by whether it met the outcome threshold. For intervention outcomes, positive cases were constructed with the initiation of interventions, and negative cases were sampled from control interventions, so that models could not rely on autocorrelation between past and future interventions. For all models, trajectories were truncated 5 minutes before the outcome and at the most recent 512 tokens.

MINT was compared against task-specific models: XGBoost trained on bag-of-words features and a regularized logistic regression trained on triage details. Differences in the AUROC were estimated with visit-paired bootstrapping and considered important when the 95% confidence interval did not include zero. We evaluated MINT against a Delphi-2M-style^5^ baseline where data is tokenized at the day level (“daily” in **Extended Data Figure 1**), and against ETHOS for only the hospital admission outcome^11^. We are unable to compare against additional EHR foundation models, as none produce age-adjusted pediatric predictions at the minute-scale.

### Multicenter validation

MINT was externally validated on pediatric ED visits from 2012 to 2026 at seven hospitals from four outside University of California health systems, accessed from the UCHDW. Whereas UCSF data were Epic-based (Verona, WI), the UCHDW represents data in the Observational Medical Outcomes Partnership (OMOP) Common Data Models. External trajectories were re-tokenized and required modified eligibility criteria, leaving more than one third of the MINT vocabulary unmapped (**Supplementary Tables 1-2**). Hospitals with sufficient event volume were evaluated individually and smaller community hospitals within a health system were pooled, yielding five analysis sites (**Figure 1c**).

The forecasting setup was otherwise identical to the internal evaluation, except that trajectories were truncated to 128 tokens for computational constraints, the minimum trajectory length was loosened to six tokens (reflecting sparse vital sign documentation in some acute care settings)^79,80^, and a few additional post-processing steps. These post-processing steps included: (1) automatically shifting forward some tokens with negative timestamps to address timestamp rounding, (2) truncating all trajectories at a maximum of 12 hours to handle incorrectly documented ED disposition times, and (3) including only visits that had tokens at three or more unique timepoints to remove for rare cases where all timestamps were rounded to midnight or noon, thereby removing the temporal component of token sequences. Evaluation was restricted to the five physiologic deterioration outcomes, because the external sites incompletely captured interventions (**Supplementary Table 2**).

At external sites, we also evaluated different permutations of MINT, including continued pre-training, transfer learning, and outcome-specific fine-tuning. For these experiments, we randomly split visits into training and test sets (50-50) using a fixed random state (seed=42) and reserved 20% of the training visits for validation. All model development and checkpoint selection were performed using the training and validation splits only, and the held-out test split was used exclusively for final evaluation. Full details on these models are available in the **Supplementary Methods**.

### Decision support for positive pressure ventilation

We compared MINT against physicians alone, physicians augmented with MINT as decision support, and two large language models. All four physicians are board-certified in emergency medicine and/or pediatric emergency medicine, and all actively practice clinically. Our experimental design followed the general framework introduced in the Lundberg et al.^34^ This task forecasted PPV initiation within 5 to 60 minutes among children with severe respiratory distress.

To assemble this cohort, we selected only children who presented with a respiratory chief complaint: respiratory distress, allergic reaction, shortness of breath, croup, wheezing, and asthma. We filtered to only windows where the outcome occurred within 60 minutes of the final non-truncated token to prevent large gaps that would impact forecasting abilities. These criteria increased PPV incidence to 14.9%, and we further upsampled to a 28% incidence to reflect that physician gestalt can rule out the need for PPV in most respiratory distress cases that are not already severe.

The final evaluation cohort included 100 visits, split across four sub-cohorts of 25 visits (7 PPV exposures each; 28%). Each of the four physician reviewers evaluated a sub-cohort with access to MINT risk estimates and SHAP force plots (augmented), and a different sub-cohort without MINT risk estimates (alone/blinded). In this design, each cohort was reviewed by two physicians, one augmented and one blinded, to evaluate the effect of model predictions on human forecasting performance.

MINT estimates were provided to physicians as isotonic regression-calibrated probabilities fit on the training set with the same inclusion criteria (**Supplementary Figure 8**), and SHAP values were provided as a force plot at the final, untruncated timepoint. Physicians were blinded to each other’s predictions and informed before the task that the outcome incidence would be 25-30%. Physicians were provided information in a review interface, based on the Prescience study^34^, that included all information provided to the model (**Supplementary Figure 9**).

The two large language models evaluated were GPT-5.5 (gpt-5.5-2026-04-24; all parameters default) and GPT-5-mini (gpt-5-mini-2025-08-07; all parameters default)^49,53^. Both LLMs system prompt asked it to predict the probability of PPV, and it was provided with 5 positive and 10 negative trajectories for in-context learning.

### Out-of-vocabulary outcomes

We trained linear probes to forecast four outcomes at two different timepoints: hospital admission and ICU admission using the first hour of ED data, and inpatient sepsis and septic shock using the first four hours of ED data. For all outcomes, visits were truncated at the earlier of the fixed timepoint and ED disposition. The admission outcome was defined as any child admitted to the hospital immediately after their ED stay. The ICU outcome was defined as any child who was admitted to the ICU at any point in their hospital visit, even if they were originally admitted to the floor. We used linear probes because the admission outcomes were measured at fixed timepoints, and because sepsis outcomes were not natively in the model’s vocabulary.

In addition to the task-specific model comparators, we reported the performance of the Emergency Severity Index (ESI) and the Emergency Department Pediatric Early Warning Score (ED-PEWS)^81,82^. ESI is a five-level triage system that scores patients on acuity and expected resource utilization; it is used in 94% of EDs across the country^81^. Visits with an unknown ESI score were excluded from the ESI evaluation. ED-PEWS is a physiologic risk score designed to identify the highest risk and lowest risk children in the ED by assigning a score from 0 (low) to 68 (high)^82^. We calculated children’s ED-PEWS score using only the first 30 minutes of data from tokenized clinical trajectories. If multiple vitals were present in this period, the highest risk ones were used to calculate the score. Visits missing a documented respiratory rate, oxygen saturation, or pulse in this period were excluded. Exam findings used in ED-PEWS, including work of breathing, decreased level of consciousness, and delayed capillary refill, were extracted from the triage information in ED physician notes with an LLM (GPT-5-nano; gpt-5-nano-2025-08-07), an approach we have previously leveraged for extracting diverse pediatric ED predictor variables^83^.

The sepsis and septic shock experiments were performed closely following the methods in Alpern et al.^54^ Children with sepsis or septic shock were defined as those meeting Phoenix Sepsis Criteria (PSC)^84^ during the first 48 hours of their inpatient stay, following the 24-hour feature window strategy in the original study. Visits with suspected infection were eligible as defined in the original study. Visits already meeting PSC criteria in the ED or discharged directly from the ED were excluded. Notably, due to data limitations, we were unable to exclude children with trauma diagnosis codes or those younger than two months old.

### Dynamic monitoring and risk estimation

We evaluated MINT’s continuous monitoring in a virtual ED simulation (**Figure 4a-b**). We randomly selected 100 children with a 20% admission rate and re-ranked them every minute by their zero-shot next-token probability of hospital admission. We repeated 1,000 simulations to identify resource-intensive visits, 30 minutes after arrival (**Figure 4c**).

To explain dynamic changes in risk, we computed SHAP (SHapley Additive exPlanations) values at every minute for selected children^34^. We calculated the change in the log-probability of hospital admission at every timestep compared to the log-probability at arrival (t=0), which typically includes age/sex, chief complaint, triage acuity, arrival method, and any immediate vitals. This approach provides more effective patient-specific tracking than a global log-probability baseline shared across all children, since children with high-risk triage details (e.g., arrival by helicopter) would always be significantly above a global baseline.

At each evaluation time point, the full token sequence up to that minute was provided to SHAP’s explainer, which systematically masks tokens from the input and measures how each token’s presence or absence changes the model’s risk estimate^34^. Individual token contributions were aggregated into feature groups for visualization as a stacked force plot over time, and representative token categories (horizontal polygons) and clinical events (vertical lines) were annotated.

### Evaluating physiologic associations and individual responses

To evaluate physiologic associations, we performed population-level counterfactuals on visits with Immediate (ESI 1) or Emergent (ESI 2) triage acuity levels (n=12,885 visits). Medication and procedure outcomes were evaluated on trajectories truncated at the earlier of ED disposition or 120 minutes after arrival. For epinephrine, transfusion, and naloxone, a single token was added to the end of the trajectory. Epinephrine was also tested with a higher dose, simulated as three tokens, each five minutes apart. Rapid sequence intubation was simulated as the sequential administration of ketamine, rocuronium, and propofol, each one minute apart. Massive transfusion protocol was simulated as blood product transfusions every five minutes for 30 minutes (six total transfusions), as well as calcium chloride and calcium gluconate with the first transfusion and tranexamic acid with the first and sixth transfusions; this analysis was performed on a subset of visits with injury chief complaints. Lastly, respiratory support was tested via token removal, rather than addition, in the subset of children who had hypoxia and later received respiratory support (n=1,378 visits). Rather than using a fixed timepoint, the intervention condition for respiratory support was evaluated immediately after the first administration of oxygen, and the baseline was the prediction immediately before the respiratory support was initiated.

For individual-level vital signs regression, we identified visits that contained a triplet sequence of a vital sign (before), medication, and a vital sign (after) occurring within 30 minutes; we selected only the subset of these windows where no other medication or procedure was administered in the 30-minute period to limit the physiologic influence of other interventions. We selected three types of token triplets for evaluation: pulse with epinephrine, oxygen saturation with albuterol, and systolic blood pressure with ketamine. For each triplet, MINT evaluated the sequence truncated immediately after the medication administration and regressed an expected post-medication vital prediction via a weighted softmax over the logits for that vital sign type (zero-shot). True versus predicted deltas were compared with scatter plots and R^2^. We also characterized the learned token embedding space with UMAP projection and vector analogies (**Figure 1b**, **Extended Data Figure 5**).

### Hypothesis generation via counterfactual simulations

To evaluate hypothesis generation capabilities, we performed counterfactuals on visits closely following inclusion criteria from two recent registry-scale studies. We evaluated whether counterfactuals generated by MINT could recover the direction of treatment-outcome associations reported in two recent registry-scale studies (**Table 1**). For ondansetron, we sampled 1,000 visits among children 10 years old or younger with chief complaints of emesis, diarrhea, or dehydration and ondansetron administration between 10 and 120 minutes after arrival. For early analgesia, we included visits with a chief complaint of sickle cell pain crisis and analgesic administration within the first 60 minutes. Each trajectory was truncated immediately after the first qualifying treatment. Counterfactual sequences were constructed by removing ondansetron tokens or delaying the first qualifying analgesic administration by 60 minutes, while retaining the remaining observed context. We repeated inference for each modified sequence and summarized within-visit changes in predicted admission probability as the proportion of visits with increased probability of admission.

## Data Availability

The University of California does not permit the public release of the de-identified data used in this study. Code will be made publicly available upon peer-reviewed publication of this study, and model weights will be distributed consistent with institutional and regulatory policy.

## Acknowledgements

The authors acknowledge the use of the University of California, San Francisco (UCSF), Information Commons computational research platform, developed and supported by UCSF Bakar Computational Health Sciences Institute. The authors thank the Center for Data-driven Insights and Innovation at UC Health (CDI2; https://www.ucop.edu/uc-health/departments/center-for-data-driven-insights-and-innovations-cdi2.html), for its analytical and technical support related to use of the UC Health Data Warehouse and related data assets.

---

**Extended Data Figure 1.**
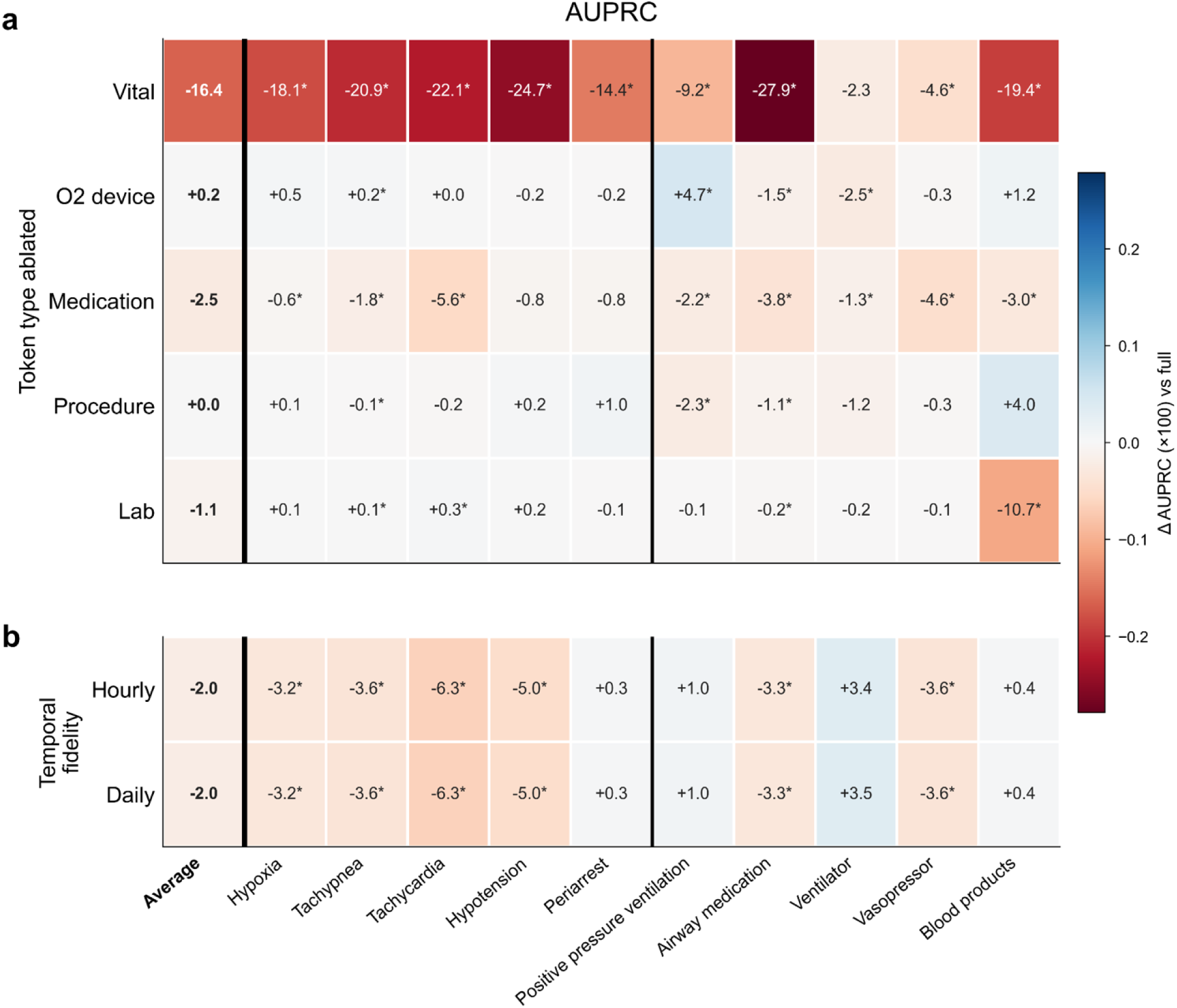
Ablations on tokenization categories and timescale. Impact of ablations on the AUPRC of zero-shot MINT forecasts, compared against the full MINT model. **Panel A**, inference time ablations on token types. **Panel B**, training and inference time ablations on token temporal fidelity (e.g. token times rounded to the nearest hour or day, instead of minute-scale).

**Extended Data Figure 2.**
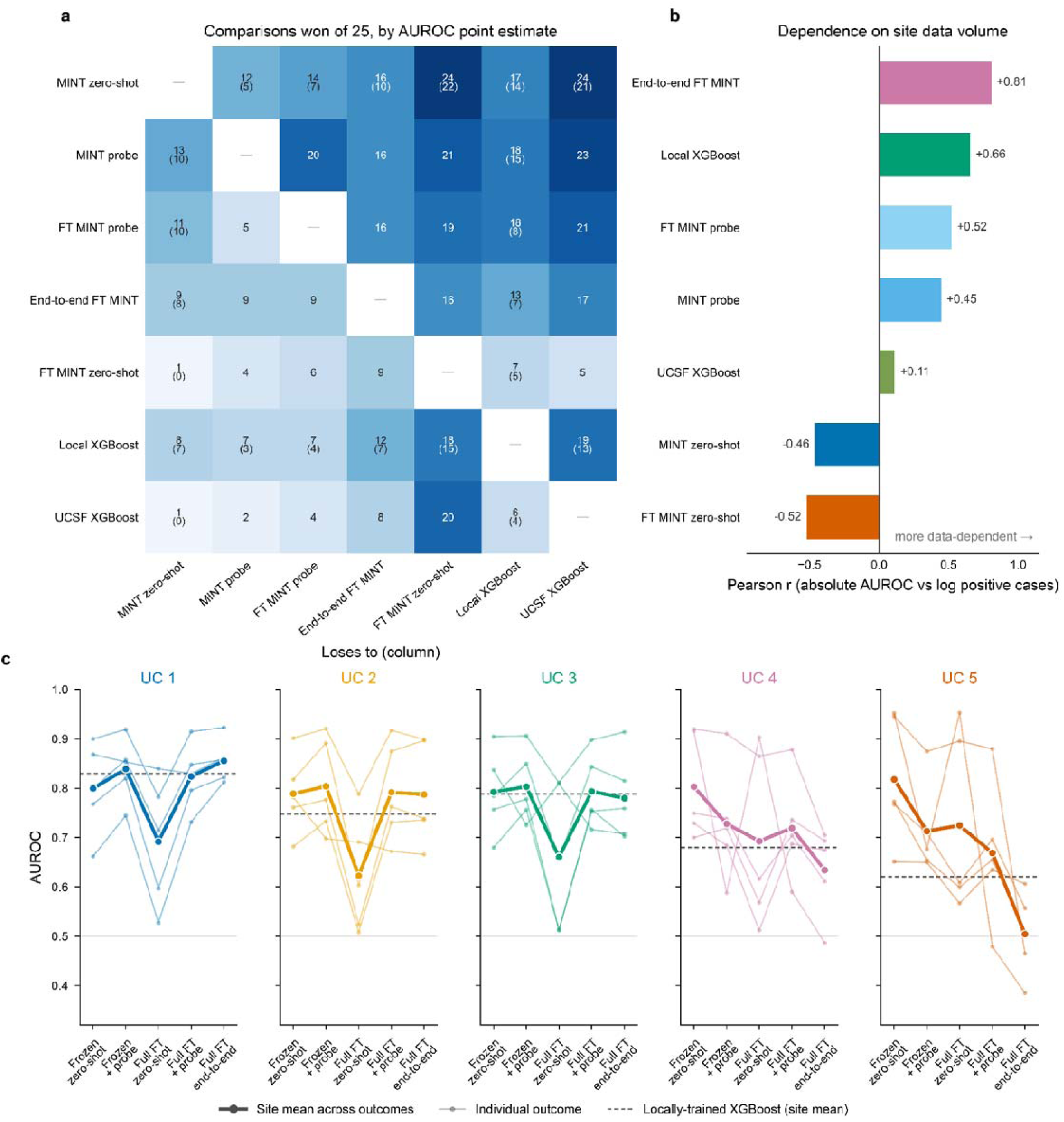
Additional findings across external generalization. **Panel A**, pairwise wins and losses across the 25 site-outcome comparisons. Each cell gives the number of comparisons in which the row method achieved a higher AUROC point estimate than the column method; values in parentheses give the number in which the 95% confidence interval of the paired difference excluded zero. Paired bootstrap estimates were available only against the zero-shot MINT and site-specific XGBoost references, so parenthetical counts appear in those two rows and columns only. **Panel B**, Pearson correlation between each method’s absolute AUROC and the number of positive cases available in that site-outcome comparison. **Panel C**, AUROC under varying degrees of site-specific adaptation, from left to right: frozen MINT zero-shot, a linear probe on frozen MINT, MINT fine-tuned on site data, a linear probe on fine-tuned MINT, and end-to-end fine-tuning with a classification head. Thin lines show individual outcomes, thick lines the mean across the five outcomes at that site, and dashed lines the mean AUROC of the site-specific XGBoost model at that site.

**Extended Data Figure 3.**
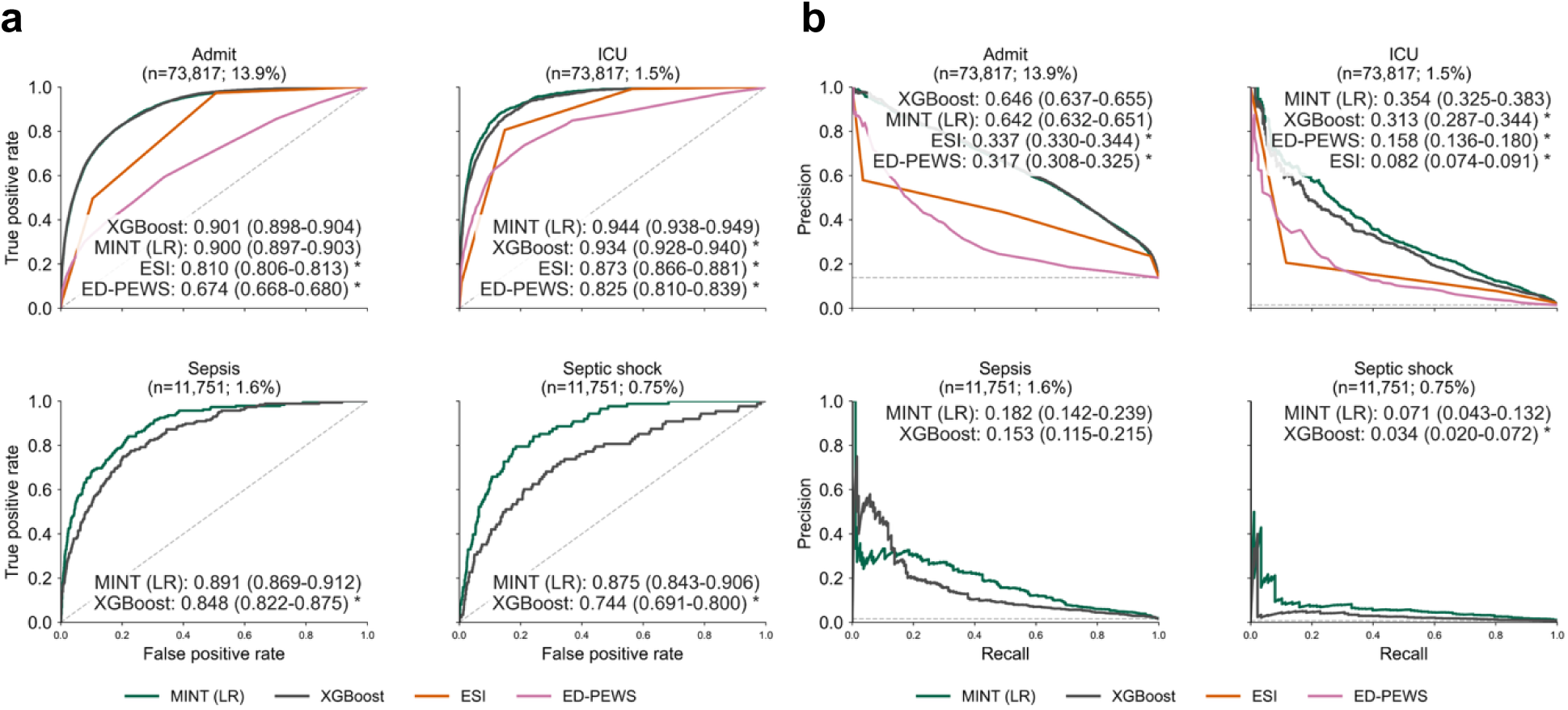
Out-of-vocabulary forecasts with MINT linear probes. Hour-scale outcomes forecasted using logistic regression probes trained on MINT representations (MINT [LR]) and compared against XGBoost. **Panel A**, performance reported with AUROC. **Panel B**, performance reported with AUPRC. Admission models were also compared to Emergency Severity Index (ESI) and Emergency Department Pediatric Early Warning Score (ED-PEWS).

**Extended Data Figure 4.**
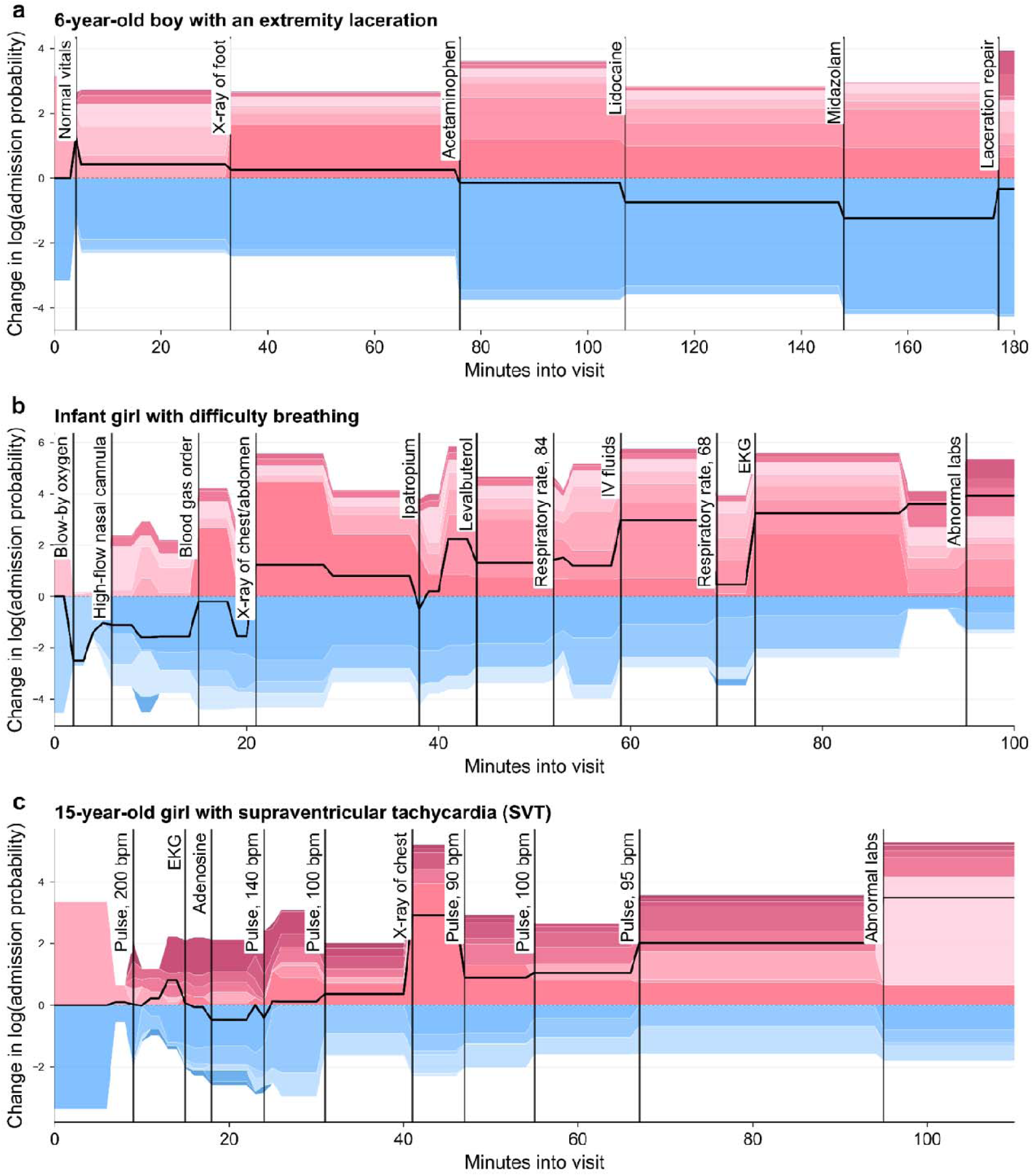
Representative temporal risk trajectories and dynamic explanations for individual children. Temporal risk plots, showing the change in log probability of admission compared to triage, for three children of varying acuity. Stacked regions represent positive (in red) and negative (in blue) token contributions, and different shades are different token categories. Key clinical events are marked with vertical lines.

**Extended Data Figure 5.**
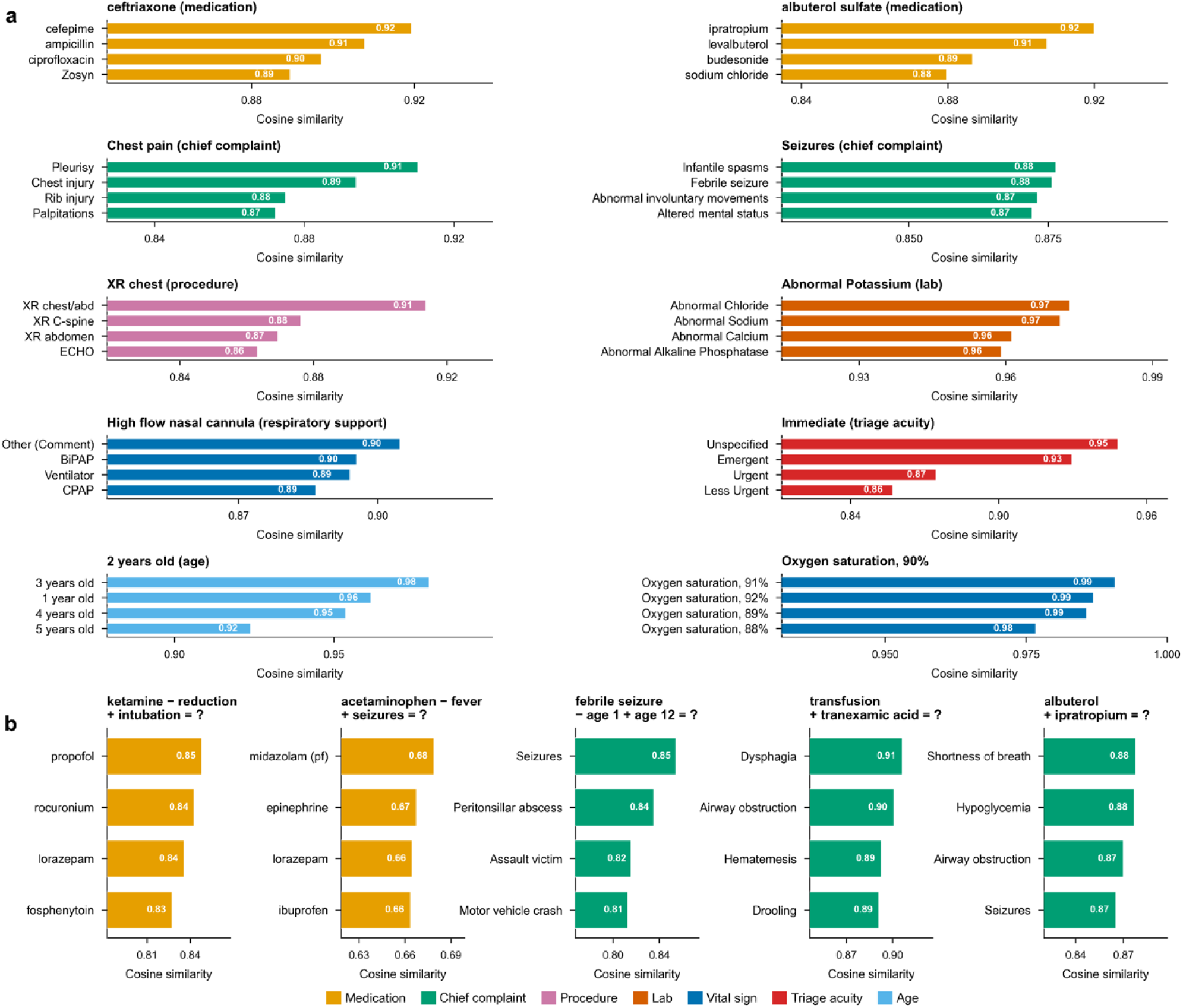
Semantic organization in the MINT token space. MINT token embeddings cluster with clinical relevance. **Panel A**, nearest neighbor identification for representative tokens, ranked by cosine similarity in the learned embedding space. **Panel B**, vector arithmetic in the token embedding space demonstrated clinically meaningful transformations across token types.

**Extended Data Table 1.** Characteristics of the visits at UCSF and external UC health systems. In external UC health systems, race categories are not mutually exclusive, and some visits with more than one recorded race may appear in both.

| UCSF | Training | Evaluation |
| --- | --- | --- |
| **Total visits** | 283682 | 81285 |
| **Date range** | 2011-2023 | 2024-2025 |
| **Tokens, median [IQR]** | 21 (16-28) | 21 (16-28) |
| **Race** |  |  |
| Asian | 13.1% (37159) | 11.6% (9449) |
| Black | 18.0% (51085) | 16.7% (13554) |
| White | 22.3% (63153) | 18.6% (15083) |
| Other/Unknown | 46.6% (132285) | 53.1% (43199) |
| **Ethnicity - Hispanic or Latino** | 41.0% (116375) | 45.7% (37132) |
| **Sex - Female** | 46.0% (130479) | 45.5% (37011) |
| **Age, median [IQR]** | 5 (1-10) | 5 (2-10) |

|  | UC 1 | UC 2 | UC 3 | UC 4 | UC 5 |
| --- | --- | --- | --- | --- | --- |
| **Total visits** | 212654 | 85790 | 73403 | 19428 | 10491 |
| **Date range** | 2012-2026 | 2013-2026 | 2013-2026 | 2017-2026 | 2012-2026 |
| **Tokens, median [IQR]** | 23 (17-35) | 17 (13-25) | 18 (15-28) | 16 (11-23) | 15 (11-20) |
| **Race** |  |  |  |  |  |
| Asian | 7.0% (14987) | 4.6% (3966) | 7.2% (5263) | 9.8% (1901) | 7.2% (752) |
| Black | 15.1% (32205) | 7.8% (6687) | 9.4% (6924) | 3.8% (735) | 11.8% (1239) |
| White | 29.9% (63647) | 49.6% (42518) | 45.2% (33199) | 53.4% (10376) | 41.7% (4371) |
| Other/Unknown | 53.3% (113259) | 47.2% (40456) | 47.2% (34616) | 33.7% (6546) | 42.4% (4446) |
| **Ethnicity - Hispanic or Latino** | 35.0% (74400) | 39.3% (33707) | 36.7% (26972) | 55.5% (10792) | 30.7% (3223) |
| **Sex - Female** | 45.0% (95688) | 45.0% (38623) | 47.2% (34656) | 47.4% (9216) | 48.3% (5072) |
| **Age, median [IQR]** | 6 (2-12) | 6 (2-12) | 7 (2-13) | 10 (3-15) | 9 (3-15) |

**Extended Data Table 2.** Definitions of minute-scale outcomes. *There is a well-established body of literature indicating that high-flow nasal cannula delivers positive pressure, particularly in children (<u>Vieira</u>, <u>Sharma</u>, <u>Harris</u>, <u>Nagler</u>).

| Outcome | Definition |
| --- | --- |
| Hypoxia | Oxygen saturation <93% |
| Tachypnea | Above Pediatric Advanced Life Support (PALS) age-specified thresholds |
| Periarrest | Pulse <60 beats per minute (bpm) in children aged 0-12 yo; adapted from PALS |
| Tachycardia | Pulse ≥220 bpm in 0 yo or ≥180 bpm in 1-12 yo; adapted from PALS |
| Hypotension | Below PALS age-specified thresholds |
| Invasive mechanical ventilation | Mechanical ventilation (ventilator) |
| Any positive pressure ventilation | Any of the following respiratory support: high-flow nasal cannula (HFNC),* continuous positive airway pressure (CPAP), bilevel positive airway pressure (BiPAP), bag-valve mask, or mechanical ventilation (ventilator) |
| Airway rescue medications | Any of the following medications: albuterol, ipratropium, racepinephrine, dexamethasone, magnesium sulfate; list adapted from UCSF respiratory clinical pathways (UCSF) |
| Vasopressors | Any of the following medications: epinephrine, norepinephrine, dopamine, dobutamine, milrinone; list adapted from the Phoenix Sepsis Criteria (Sanchez-Pinto) |
| Blood product transfusion | Any blood product orderable |

**Extended Data Table 3.** Extended performance for forecasting positive pressure ventilation by MINT, two large language models, and physicians. MINT cost was calculated via the energy usage to run 100 cases on a M2 MacBook Pro to simulate use at community sites without dedicated GPU hardware. The total energy usage was 2.5635e-05 kilowatt-hour (kWh), estimated with CodeCarbon (<u>CodeCarbon</u>). Assuming average energy costs in the United States at the time of writing ($0.19 USD/kWh), the runtime costs were $4.87e-06. Runtime was calculated with no parallelization for any method.

| Method | Cost to run<br>(100 cases;<br>\$USD) | Runtime<br>(100 cases;<br>seconds) | AUROC | AUROC<br>difference vs<br>MINT | AUPRC | AUPRC<br>difference vs<br>MINT |
| --- | --- | --- | --- | --- | --- | --- |
| **MINT** | \$4.87e-06 | 0.99 seconds | 0.903<br>(0.827 to<br>0.962) | - | 0.794<br>(0.678 to<br>0.925) | - |
| **Physicians**<br>**with MINT** | Not estimated | Not<br>estimated | 0.855<br>(0.764 to<br>0.926) | -0.048 (-0.100<br>to 0.002) | 0.744<br>(0.613 to<br>0.865) | -0.050 (-0.145<br>to 0.028) |
| **Physicians**<br>**alone** | Not estimated | Not<br>estimated | 0.784<br>(0.679 to<br>0.875) | -0.119 (-0.206<br>to -0.035) | 0.655<br>(0.505 to<br>0.816) | -0.139 (-0.298<br>to 0.006) |
| **GPT-5.5** | \$3.70 | 830 seconds | 0.863<br>(0.774 to<br>0.936) | -0.040 (-0.101<br>to 0.017) | 0.735<br>(0.594 to<br>0.873) | -0.058 (-0.181<br>to 0.057) |
| **GPT-5-mini** | \$0.22 | 730 seconds | 0.784<br>(0.687 to<br>0.875) | -0.119 (-0.205<br>to -0.041) | 0.619<br>(0.469 to<br>0.775) | -0.175 (-0.326<br>to -0.030) |

## Notes

Funding Information: This work is supported by the Eunice Kennedy Shriver National Institute of Child Health and Human Development of the National Institutes of Health under award number K23HD110716 (AK). This information or content and conclusions are those of the authors and should not be construed as the official position or policy of, nor should any endorsements be inferred by HRSA, HHS or the U.S. Government.

### Competing Interest Statement

A.K. and N.A. are co-founders and consultants to CaptureDx. This entity had no role in the design, planning, or execution of the study, or interpretation of the findings. The other authors do not have competing interests.

### Author Declarations

The Institutional Review Board of the University of California, San Francisco waived ethical approval for this work.

