## Supplementary Materials for "Learning the electronic health record at the minute-scale"

Narang et al. 2026

**Supplementary Methods**

*Additional details on model training and adaptations from Delphi-2M*

We made minimal changes to the Delphi-2M architecture ([Shmatko](https://www.nature.com/articles/s41586-025-09529-3)) in the development of MINT. First, we updated the vocabulary size to 1,475 (plus special tokens) to reflect our tokenization strategy. Second, we reduced the token timescale from days since birth to minutes since emergency department (ED) arrival. Third, we disabled lifestyle augmentations, since they did not apply in our acute care token space.

Fourth, we performed two phases of hyperparameter optimization on our validation data (a 10% split of the dataset described elsewhere as the training set). The optimal parameters were selected by evaluating the mean area under the precision-recall curve (AUPRC) of physiologic monitoring forecasts on the validation dataset. In the first phase, we searched over block size (48 [default], 64, 96), data loading strategy (left [default], start, random), and no event rate (0, 5 [default], 60). In the second phase, we searched over token dropout (0.0 [default], 0.1) and no event rate (0.16, 5 [default]). The only change that improved validation performance was updating the block size to 64; all other parameters were kept at defaults from the original Delphi-2M study ([Shmatko](https://www.nature.com/articles/s41586-025-09529-3)).

We did not evaluate increasing the number of attention heads and layers in the model, because we aimed to keep the computational requirements low enough to enable model inference on consumer-grade laptops to enable generalizability to under-resourced centers. We confirmed that all analyses ran on a M2 Macbook Pro.

As in the original Delphi-2M study, MINT was trained with a batch size of 128 using the AdamW optimizer (β₁ = 0.9, β₂ = 0.99, weight decay = 0.2) with a peak learning rate of 6 × 10^-4^ and cosine decay to a minimum of 6 × 10^-5^ over 100,000 iterations, with a 1,000-iteration linear warmup; the model was trained for 100,000 iterations total. The composite loss function combines cross-entropy loss over next-token predictions (ignoring padding tokens) and exponential log-likelihood loss over inter-event time predictions. Validation loss was evaluated every 250 iterations using 25 batches, and the checkpoint with the lowest validation loss was retained.

*Rationale for multicenter transfer design*

A key strength of our study was testing the model’s ability to generalize across database setups and EHR systems using two of the most widely adopted data models in clinical care. The UCSF dataset was accessed from a database schema built on top of Epic’s EHR data model. Epic is the largest EHR vendor for academic centers in the United States, and the UCSF schema should be easy to replicate in other large research datasets, including Epic COSMOS data and Kaiser Permanente HealthConnect. Afterwards, the multicenter dataset was accessed from the University of California Health Clinical Data Warehouse using the Observational Medical Outcomes Partner (OMOP) Common Data Model, a widely adopted open standard for managing large EHR datasets.

In the external cohort we evaluated only physiologic outcomes, and not clinical interventions, for two reasons. First, as shown in **Supplementary Table 2**, token mapping was poor for respiratory intervention devices, meaning those interventions were not fully captured. Second, interventions are a function of local clinical practice rather than vital signs which more closely represent physiologic truth about patients. Because our external dataset includes several low-volume, non-children’s hospitals, possible discrepancies between model forecasts of interventions and actual interventions may reflect differences in clinical practice at the external hospitals rather than errors by the model. These inaccuracies would incorrectly penalize the model for performing a clinically-useful prognostic task akin to “recommending” an intervention.

*MINT zero-shot estimators*

MINT is based on the Delphi-2M architecture which introduced a novel approach of competing exponentials to forecast both the probability and time horizon of future events. The output logits are transformed into event rates by the given function λ = exp(logit) from which the probability of an event occurring within 5 minutes is calculated as P(t < 5) = 1 − exp(−exp(logit) × 5). This formulation works well when rates are constant in time which is likely less true of emergency department (ED) data than of the lifespan-scale data in the original paper, even with a well-calibrated “no event” token rate. As an alternative to exponential waiting times, applying softmax over the logits directly forecasts the token most likely to occur next.

In validation experiments, we evaluated both softmax and exponential waiting time approaches and found that softmax generally outperformed exponentials, though performance was similar. We also experimented with test-time inference variants of both techniques, modeled as sampling one step forward with the 20 most-likely next tokens from an initial trajectory and pooling zero-shot estimator probabilities across those 20 new trajectories. Though this approach did slightly improve performance for some outcomes, we decided that that 20-fold increase in inference time was not worth the relatively small improvement. However, test-time inference remains an important aim for future work.

Ultimately, we selected softmax for all downstream analyses in the study, except for the visit-level temporal plots in **Figure 4d** and **Extended Data Figure 4**. In these evaluations, we used the exponential estimator for admission, since it was better calibrated for SHAP-style explainability whereas softmax made risk estimates less interpretable.

*Bag-of-words XGBoost, linear probes, and triage logistic regression models*

The XGBoost comparator for all experiments was a gradient-boosted tree trained on the bag-of-words form of the token input, following the same truncation strategy as the MINT model for each task. All XGBoost models were fit at a fixed random state (seed=42), a binary logistic objective, and balanced class weights, with all other parameters set at defaults in XGBoost version 3.2.0.

During experiments on the validation set, we benchmarked different linear probes based on support vector machines (SVM), gradient-boosted trees (XGBoost), and logistic regression (LR). We selected LR for all linear probe experiments in this study as the simplest model that was generally the highest performing. For all linear probe experiments, LR was configured with maximum iterations of 2,000, balanced class weights, a fixed random state (seed=42), and all other parameters at defaults in scikit-learn version 1.7.2.

The triage model was designed to improve upon the demographic baselines common in foundation model literature. Recognizing that minute-scale physiology would be difficult to predict with just demographic details (age and sex), we also included ESI acuity level, chief complaint, arrival mode, and first 30 minute vital signs, roughly replicating the information known at ED triage. The bag-of-words features were used as input for a LR model with identical hyperparameters to the linear probes described above.

*Multicenter fine-tuning experiments*

For each hospital, we randomly split visits 50%-50% into training and test sets using a fixed random state (seed=42), and then reserved 20% of the training visits for validation. All model development and checkpoint selection were performed using the training and validation splits only, and the held-out test split was used exclusively for final evaluation. We used the same outcome labeling scheme as in the main analysis.

**Fine-tuned MINT** (site-level): To adapt MINT to each hospital, we fine-tuned the pretrained backbone using the original self-supervised MINT objective on site-specific trajectories. Optimization used AdamW with a target peak learning rate of 3 × 10^-5^, weight decay of 0.1, betas of (0.9, 0.99), gradient accumulation over 4 micro batches of size 8, linear warmup for 50 optimization steps, cosine decay thereafter, and gradient clipping at a norm of 1.0. We used a dropout of 0.2 during fine-tuning, evaluated the validation loss regularly, and retained the checkpoint with the lowest validation loss. Forecasts were made using zero-shot softmax, as in the UCSF-trained MINT checkpoint.

**Linear probes on MINT and fine-tuned MINT** (site-level and outcome-level): We used the linear probe approach described above, trained and evaluated separately for the original UCSF-trained MINT checkpoint and for the locally fine-tuned MINT checkpoint.

**Fine-tuned end-to-end MINT** (site-level and outcome-level): We attached a single linear output layer to the last hidden state of the UCSF-trained MINT backbone and fine-tuned the backbone and head jointly on labeled encounters. The sequence embedding was taken from the final non-padded token, mapped through a one-unit linear layer, and trained with binary cross-entropy with logits. To account for class imbalance, we used a positive-class weight equal to the ratio of negative to positive training examples. The optimization and hyperparameters were similar to the fine-tuned MINT approach described above.

**Supplementary Figures and Tables**

**Supplementary Table 1. Tokenization strategy at UCSF**

| **Token category** | **Number of tokens in category** | **Tokenization Details** |
| --- | --- | --- |
| Vitals | 578 | Vitals were selected directly from the value in the flowsheet. Vitals received the following rounding to reduce redundancy between tokens and increase training signal:   1. **Weight**: nearest integer 2. **Respiratory rate**: nearest multiple of 2 3. **Pulse**: nearest multiple of 5 4. **Oxygen saturation**: nearest integer (no rounding needed) 5. **Temperature**: nearest integer 6. **Systolic and diastolic**: nearest multiple of 2 7. **Mean arterial pressure**: nearest multiple of 5 8. **Glasgow Coma Scale**: nearest integer (no rounding needed) |
| Respiratory Support | 78 | Respiratory support devices and oxygen flow rate were selected directly from the value in the flowsheet. The flowsheet was preferred over the procedures, because flowsheet timestamps were more likely to be temporally accurate. For downstream analysis, we combined respiratory support devices into clinically grounded groupings:   1. **Room air** 2. **Blow-by oxygen** 3. **Nasal cannula** 4. **Oxygen mask** 5. **High-flow nasal cannula** 6. **CPAP, BiPAP, and BVM** 7. **Ventilator** |
| Medications | 307 | All medications with at least 300 administrations were included as tokens. To ensure that no rare rescue medications were missed, we reviewed the medications listed in Sax et al. (2024), and added five additional medications important in acute care outcomes. |
| Procedures | 93 | All procedures with at least 50 occurrences (capturing >95% of all procedures performed) were reviewed by a clinical investigator and categorized into larger categories to address redundancy and increase training signal. A total of 482 procedures were identified grouped into 93 composite categories. We excluded other procedures due to redundancy with other data sources (e.g., respiratory support was captured on flowsheets) or lack of relevance to acute care outcomes (e.g., billing details or nursing instructions). In some cases, groupings were merely semantic (e.g., EKGs were coded multiple ways), but in other cases groupings were to unify similar clinical concepts (e.g., Laceration repair of <2.5 cm and laceration repair of 2.6-5 cm both become laceration repair). |
| Laboratory Testing (abnormal) | 41 | Clinically-relevant lab tests with at least 5,000 occurrences (n=51) were identified by a clinical investigator (K.N.). Labs were included only if 5% or more of lab results for that type had an abnormal reading, excluding 10 labs. Labs were tokenized to only be included in trajectories for abnormal results (abnormality was pre-computed in the source database, accounting for age- and case-specific thresholds). Lab results that were normal were not included in any trajectories. |
| Age | 18 | Age (in years) was tokenized as an integer from 0 to 17. |
| Sex | 3 | Tokenized as female, male, or other |
| Disposition | 3 | The two primary ED dispositions were Admit and Discharge. Children with any ICU length of stay also had an ICU token added with the Admit token. |
| Chief Complaint | 328 | Chief complaints were selected directly from tabular information in the database. |
| Triage Acuity Level | 6 | Acuity was tokenized as the five Emergency Severity Index (ESI) levels directly from the EHR (Immediate, Emergent, Urgent, Less Urgent, Non-Urgent) with a sixth level for any children missing an ESI score. |
| Arrival Method | 20 | Arrival methods were selected directly from tabular information in the database. Examples include: Walk In, Car, Ambulance, Rotor Wing, Medical Flight |
| **Total** | 1,475 | There are two additional special tokens for padding (ID = 0) and no event (ID = 1); the latter concept was introduced in the original Delphi-2M study to better calibrate the time-to-event loss and forecasts. |

**Supplementary Table 2. Tokenization strategy at external health systems**

| **Token category** | **Coverage** | **Changes in tokenization** |
| --- | --- | --- |
| Vitals | 573/578 (99.1%) | Identical rounding and selection strategy to the UCSF dataset. |
| Respiratory Support | 6/78 (7.7%) | Respiratory support interventions and oxygen flow rate were poorly documented in the multicenter OMOP database. Though respiratory support interventions were technically present in the procedures table, many had timestamps rounded to noon or midnight, making them unwieldy for the minute-scale task.  The database, however, did generally have better capture of oxygen flow rates with more accurate timings in the flowsheets. We settled on a combined approach to ensure that some respiratory support signal was captured.  Oxygen flow rates were approximated to respiratory interventions with the following thresholds:   1. 0 liters per minute (lpm): Room air 2. 1-6 lpm: Nasal cannula 3. 7-15 lpm: Simple mask 4. >15 lpm: High flow nasal cannula   More intensive devices, specifically CPAP and Ventilator, were tokenized based on information in the procedure table. |
| Medications | 281/307 (91.5%) | Medications were mapped using RxNorm codes accessed from the original UCSF dataset and linked to the corresponding OMOP concept IDs in the multicenter dataset. |
| Procedures | 80/93 (86.0%) | All procedures with at least 150 occurrences across all external sites were reviewed by a clinical investigator. A total of 240 procedures were identified and mapped to the closest procedures in the UCSF-derived vocabulary. The 240 procedures mapped to 80 of the procedures in the vocabulary. |
| Laboratory Testing (abnormal) | N/A | Not available |
| Age | 18/18 (100%) | Ages may be ±1 year from the actual age, because only the year of birth was available without the month or day. |
| Sex | N/A | Available but not included, since sex was not included in the first external test run. We did not want to introduce new bias by changing the tokenization strategy after beginning evaluations on the multicenter data. |
| Disposition | N/A | Available but not accessed, since all multicenter validation occurred on data prior to ED disposition. |
| Chief Complaint | N/A | Not available |
| Triage Acuity Level | N/A | Not available |
| Arrival Method | N/A | Not available |
| **Total** | 958/1,475 (64.9%) |  |

**Supplementary Figure 1. MINT performance reported with area under the precision-recall curve (AUPRC)**

All analyses identical to **Figure 2** but with performance reported as AUPRC for minute-scale outcomes.

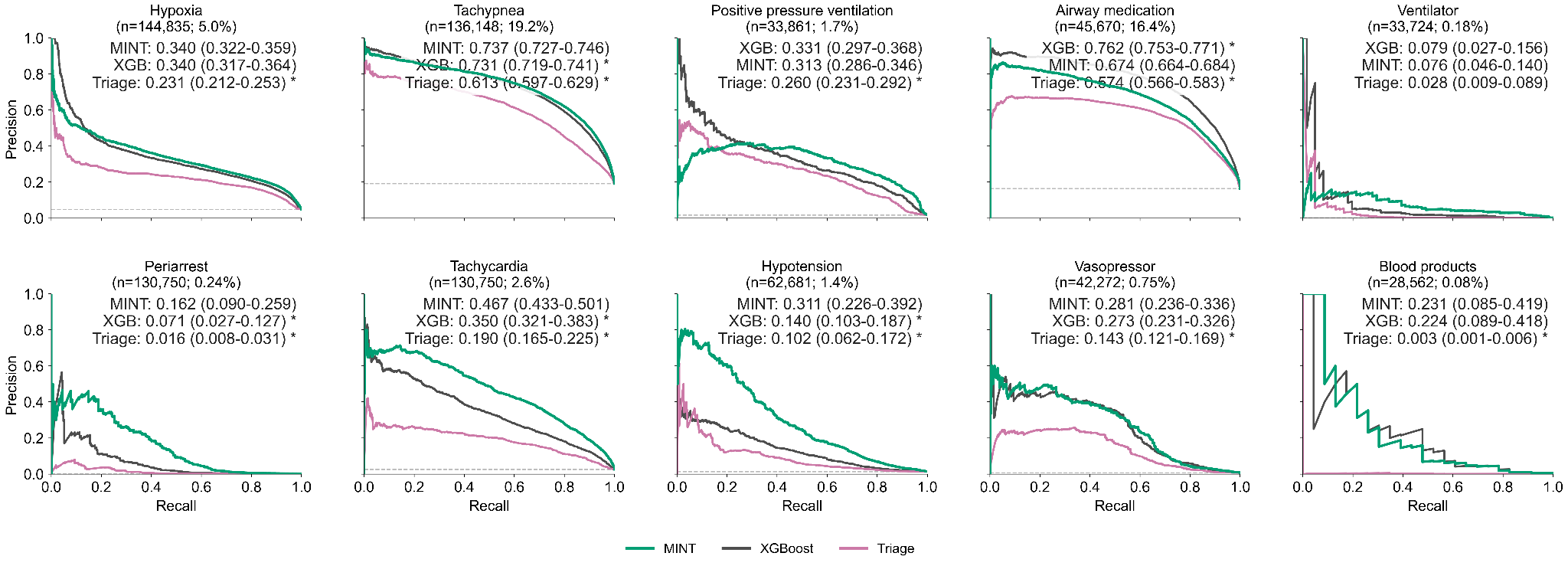

**Supplementary Figure 2. Zero-shot adaptation to varying outcome definitions**

**Panel A and B**, MINT zero-shot forecasts evaluated on multiple thresholds for hypoxia (**Panel A**) and respiratory support (**Panel B**) without additional fine-tuning or task-specific training.

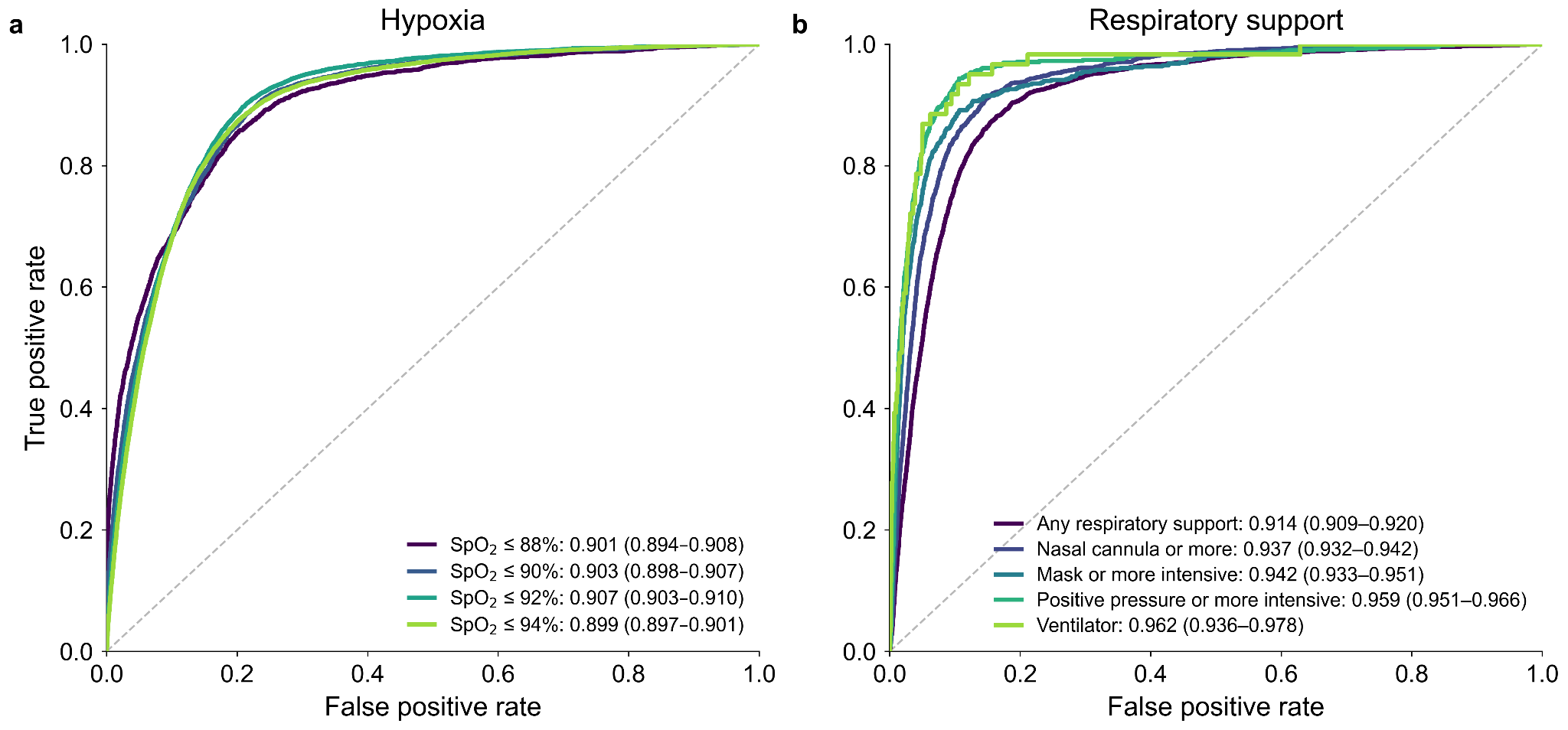

**Supplementary Figure 3. MINT representations improve sample efficiency for downstream prediction in airway medication administration**

Performance of a logistic regression (LR) probe trained on frozen MINT representations for forecasting airway medication administration compared with a task-specific XGBoost model as the amount of task-specific training data increases. The dashed horizontal line indicates the performance of the zero-shot foundation model without any task-specific training. Each point represents the mean AUROC across 10 independently trained models using the indicated number of training cases, and shaded regions denote ±1 standard deviation across model fits. The upper x-axis estimates the equivalent years of data required at a median-volume emergency department (3,870 pediatric visits per year; [Sullivan](https://pubmed.ncbi.nlm.nih.gov/23618163/)). MINT representations consistently required fewer training examples than XGBoost to achieve comparable performance and exceeded zero-shot performance with approximately 2,000 training cases (less than one year of ED data at most hospitals). This result likely underestimates the years of data required due to the higher average acuity of children presenting to UCSF’s quaternary care, academic children’s hospitals.

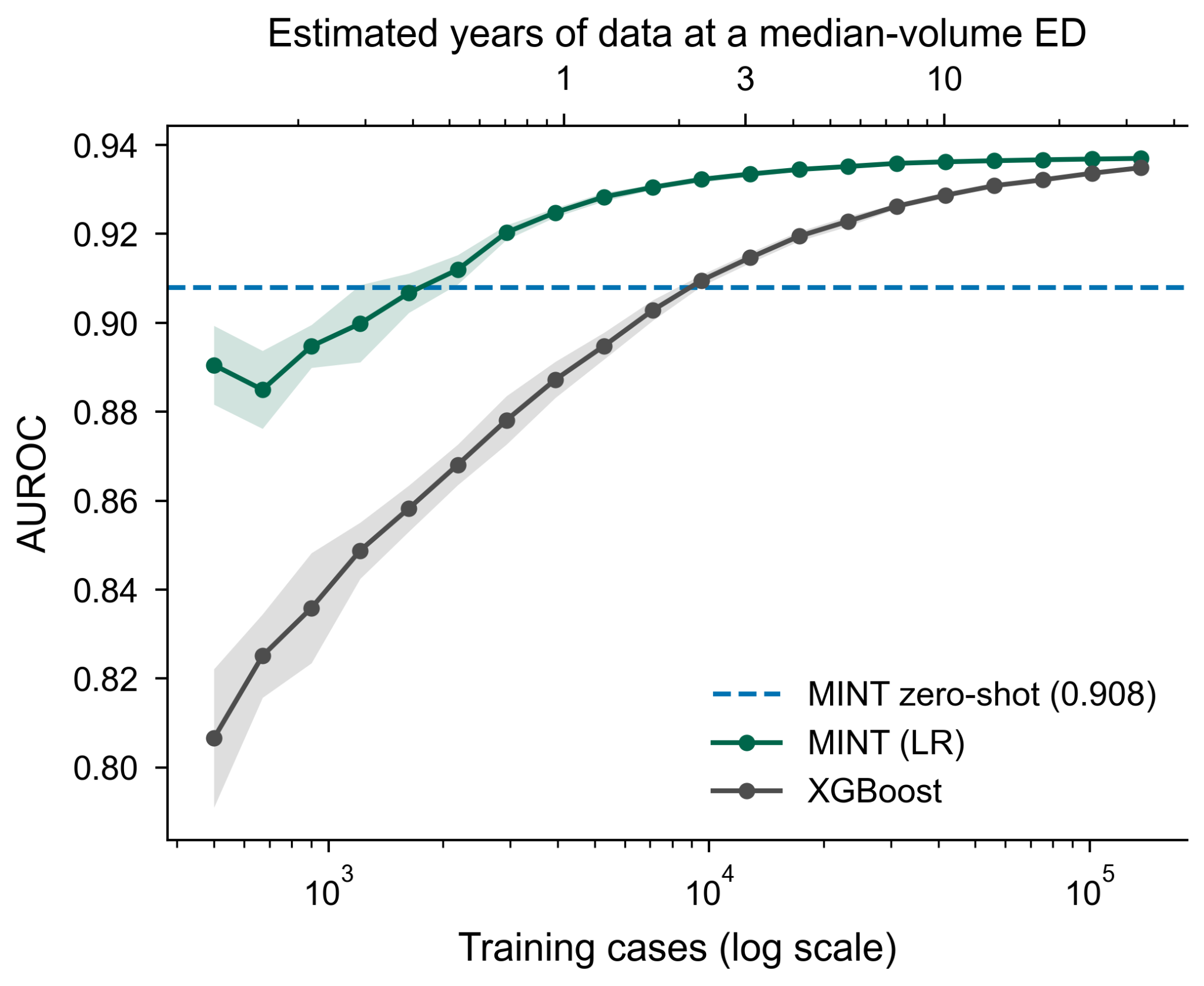

**Supplementary Figure 4. Forecasting performance for ICU admission and hypoxia stratified by demographics and insurance status**

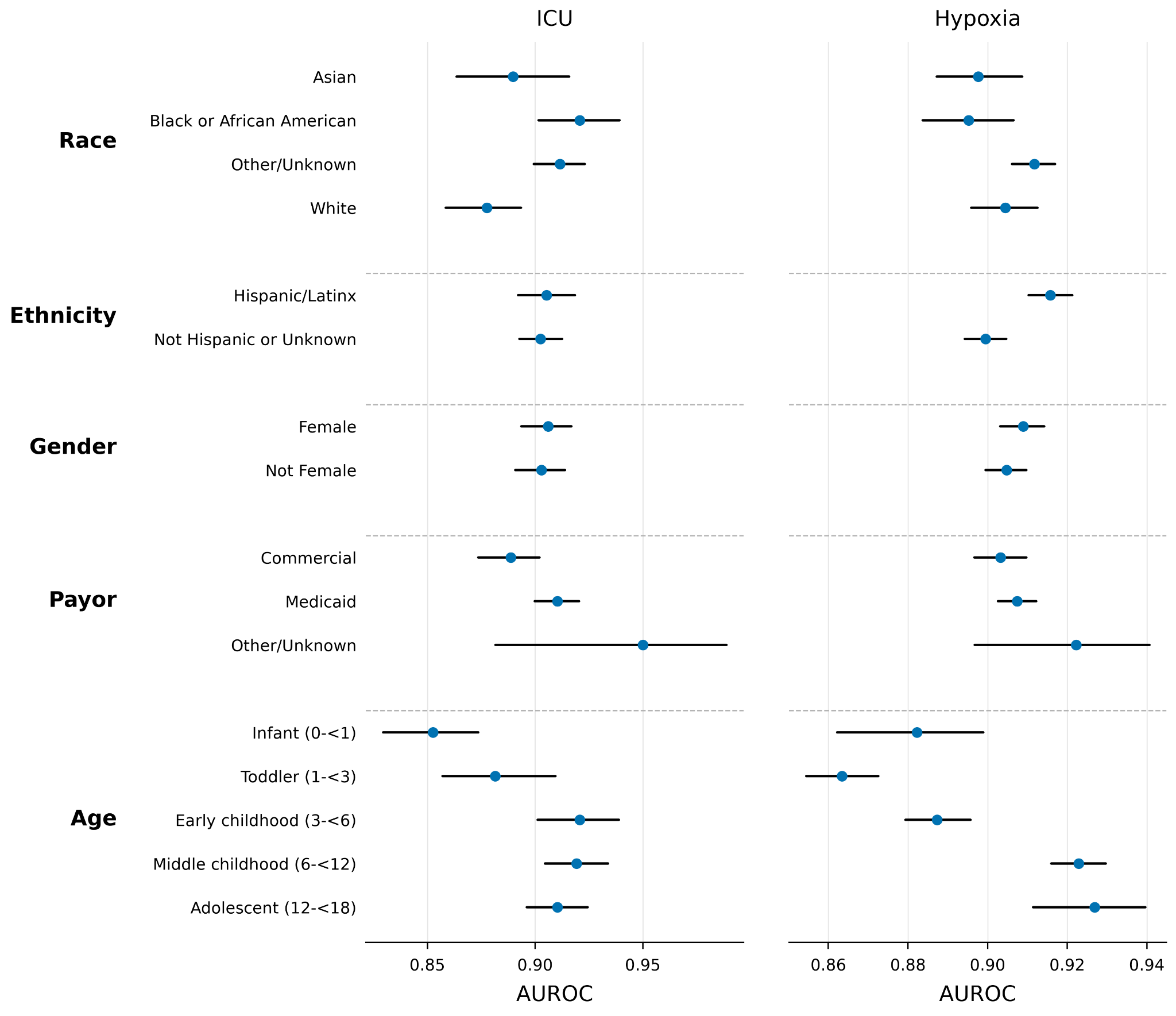

**Supplementary Figure 5. Paired differences in AUROC for all models on multicenter data**

Cells show differences in AUROC between one method and a reference for each physiologic deterioration outcome. Positive values (blue) indicate the method outperformed the reference; negative values (red) indicate the reference performed better. References are zero-shot MINT which received no site-specific or task-specific training (left) and a task-specific XGBoost model trained on site-specific data (right). Asterisks denote comparisons in which the 95% confidence interval of the difference excluded zero.

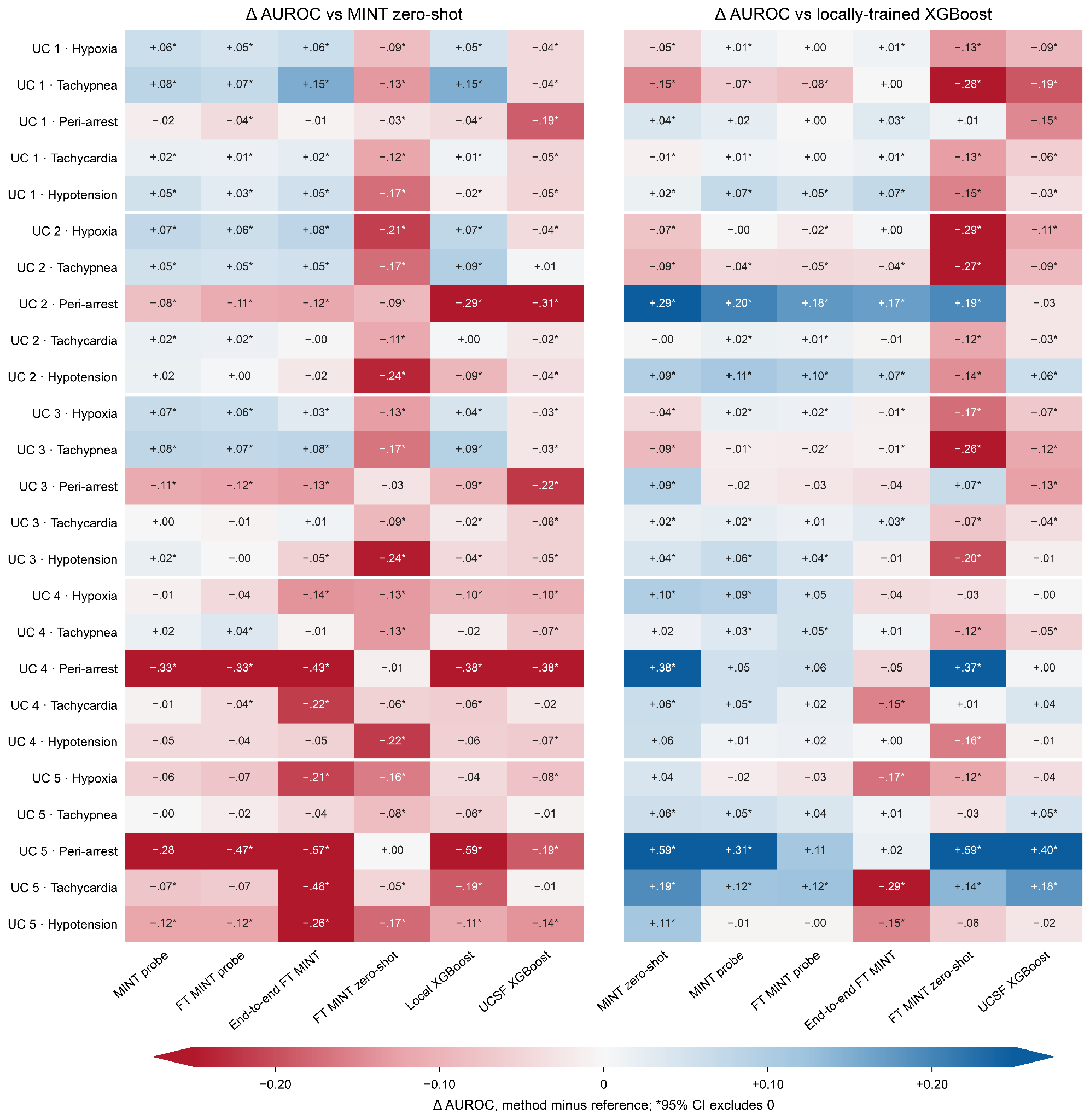

**Supplementary Table 3. ETHOS performance for forecasting hospital admission**

ETHOS performance on the held-out UCSF testing set ([Renc](https://www.nature.com/articles/s41746-024-01235-0)). Trajectories were translated to ETHOS vocabulary using an agentic workflow ([Claude Opus](https://www-cdn.anthropic.com/6a5fa276ac68b9aeb0c8b6af5fa36326e0e166dd/Claude%20Opus%204.6%20System%20Card.pdf)) supervised by a study investigator. ETHOS was tested using a logit (comparable to MINT) and rollout estimator (default approach in ETHOS). ETHOS runtime was approximately 21 times slower than MINT. Poor performance is potentially associated with pretraining only on adult data, a lack of vocabulary for important vital signs (respiratory rate, pulse, and respiratory support devices), and no evaluations on ED outcomes.

*Simulation is limited to a random subset of 2,086 visits due to computational constraints; simulations are up to 10,000x more computationally intensive than logits, requiring 20 simulations with up to 500 autoregressive forward steps per visit.

| Predicting hospital admission at 4 hours | ETHOS | |
| --- | --- | --- |
|  | **Logits** | **Simulation*** |
| **AUROC** | 0.4726 | 0.4976 |

**Supplementary Figure 6. Additional “confounding by indication” in MINT physiologic associations**

MINT demonstrated signs of confounding by indication for several important critical care medications. **Panel A**, vital sign distributions for confounded interventions. **Panel B**, mean (SD) values for selected vital signs in all children and children receiving confounded interventions.

**a**

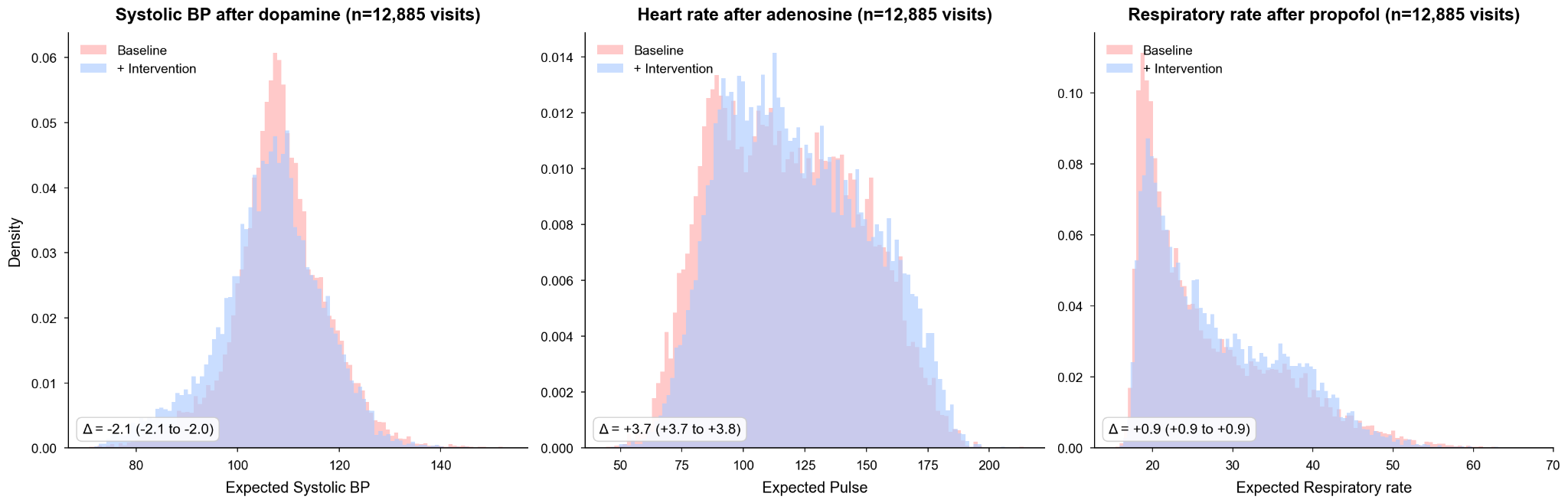

**b**

|  |  | **Mean (SD)** | |
| --- | --- | --- | --- |
| **Medication** | **Vital sign** | **All children** | **Children receiving medication**  **(pre-intervention)** |
| **Dopamine** | Systolic blood pressure (mmHg) | 110.31 (13.54) | 87.83 (12.07) |
| **Adenosine** | Pulse (beats/min) | 109.26 (27.06) | 209.27 (40.81) |
| **Propofol** | Respiratory rate (breaths/min) | 24.56 (8.00) | 24.47 (12.98) |
| **Naloxone** |  |  | 22.23 (10.14) |

**Supplementary Figure 7. Directional associations for a larger set of medications**

The directional effects of 26 medication-outcome associations were correct (blue) or incorrect (red) compared to known physiology. Bootstrapped 95% CIs were drawn but were narrow enough to be hidden behind points for most outcomes. Medication names are shown with the total number of administrations in the emergency department training set.

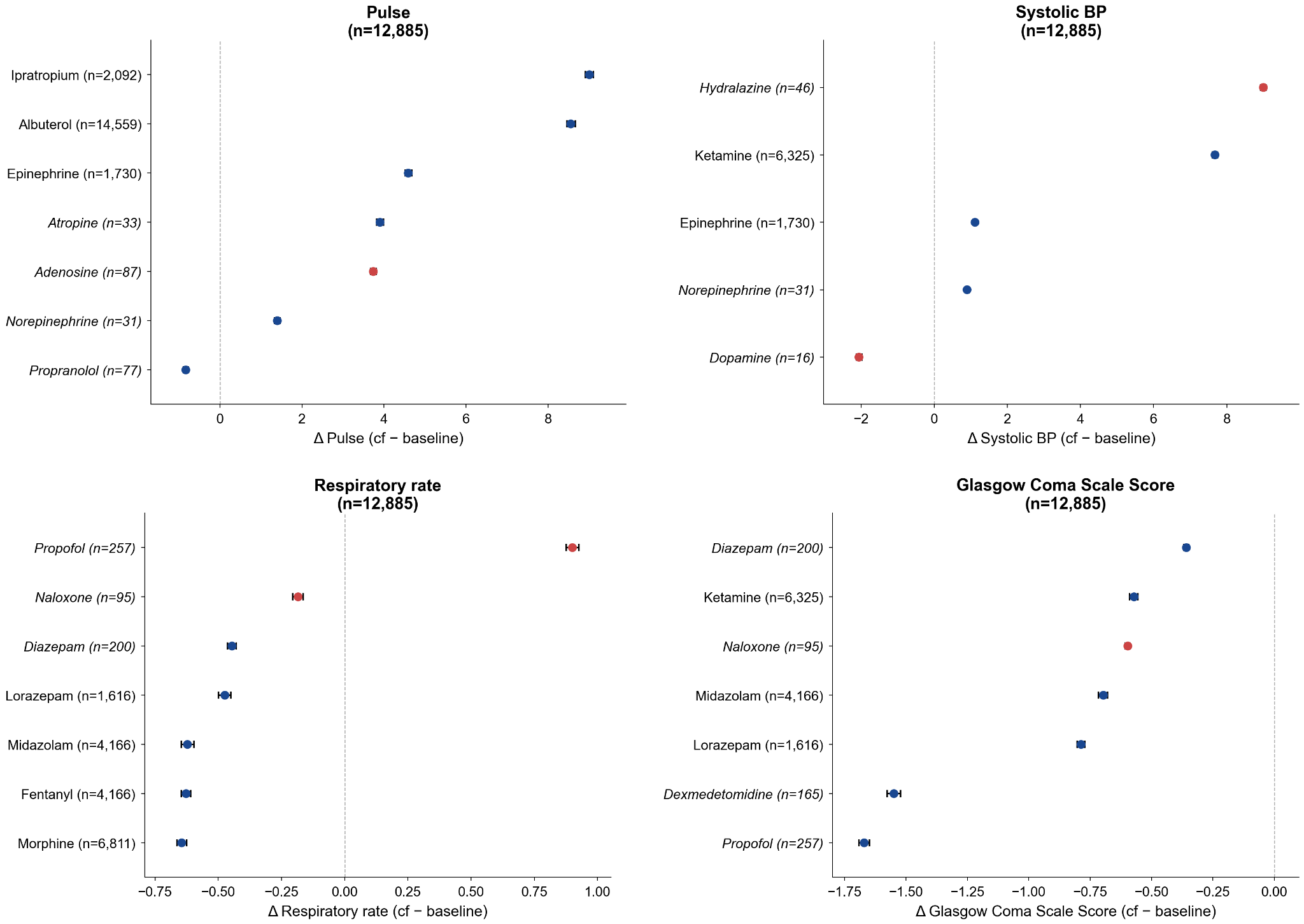

**Supplementary Figure 8. Isotonic calibration curve for MINT probability of positive pressure ventilation prior to physician reviewer interpretation**

MINT probabilities were well-calibrated after isotonic regression in a cohort of children with respiratory distress. The calibration model was fit on training data, and the shown curves are for the temporal hold-out evaluation data.

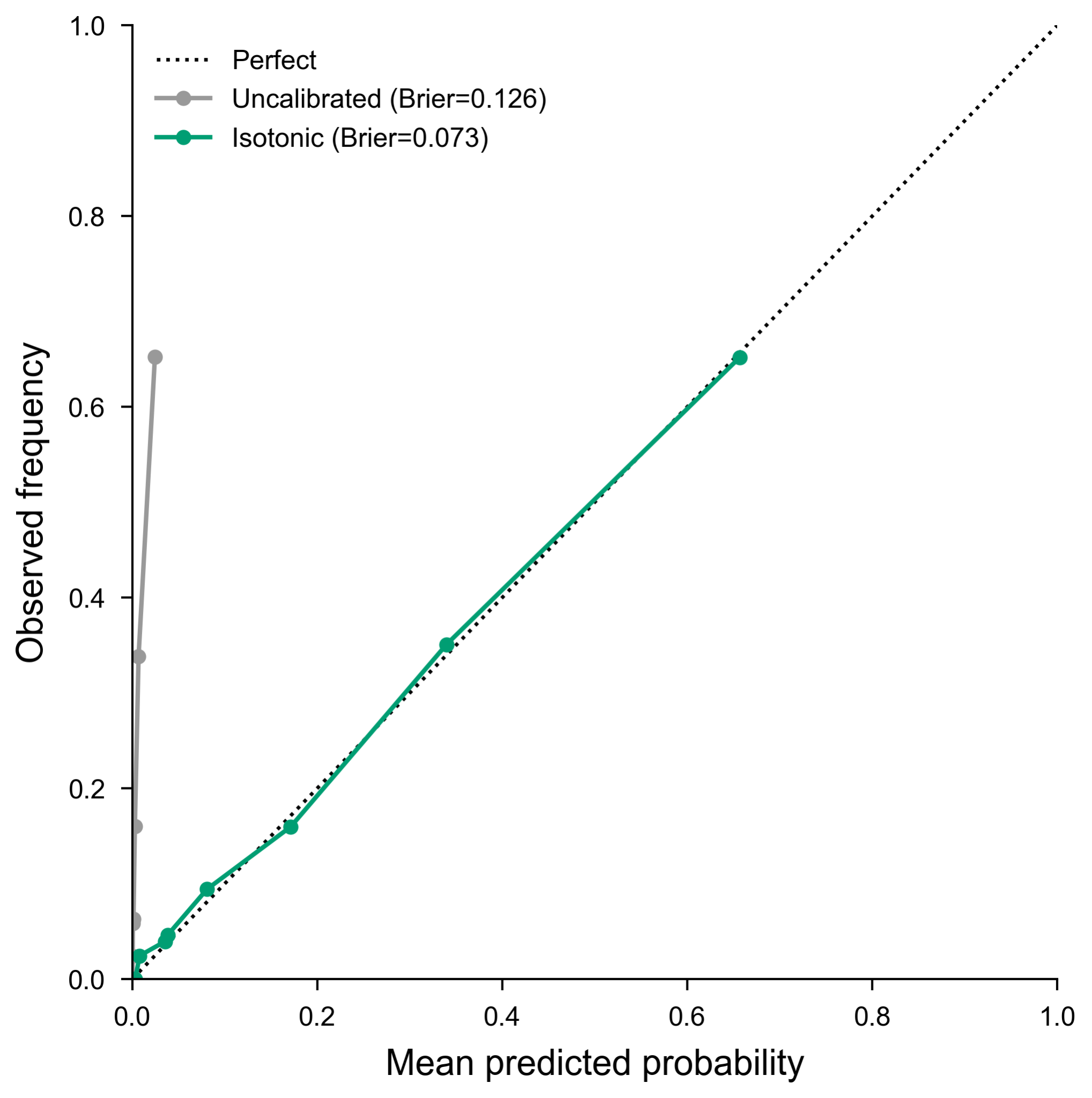

**Supplementary Figure 9. Physician review interface for positive pressure ventilation forecasting**

The review interface provided to physicians, including separate tracks for the vitals signs, medications, and procedures. Reviewers were either provided (assisted/augmented) or blinded to the MINT probability and forceplot at the top of the page.

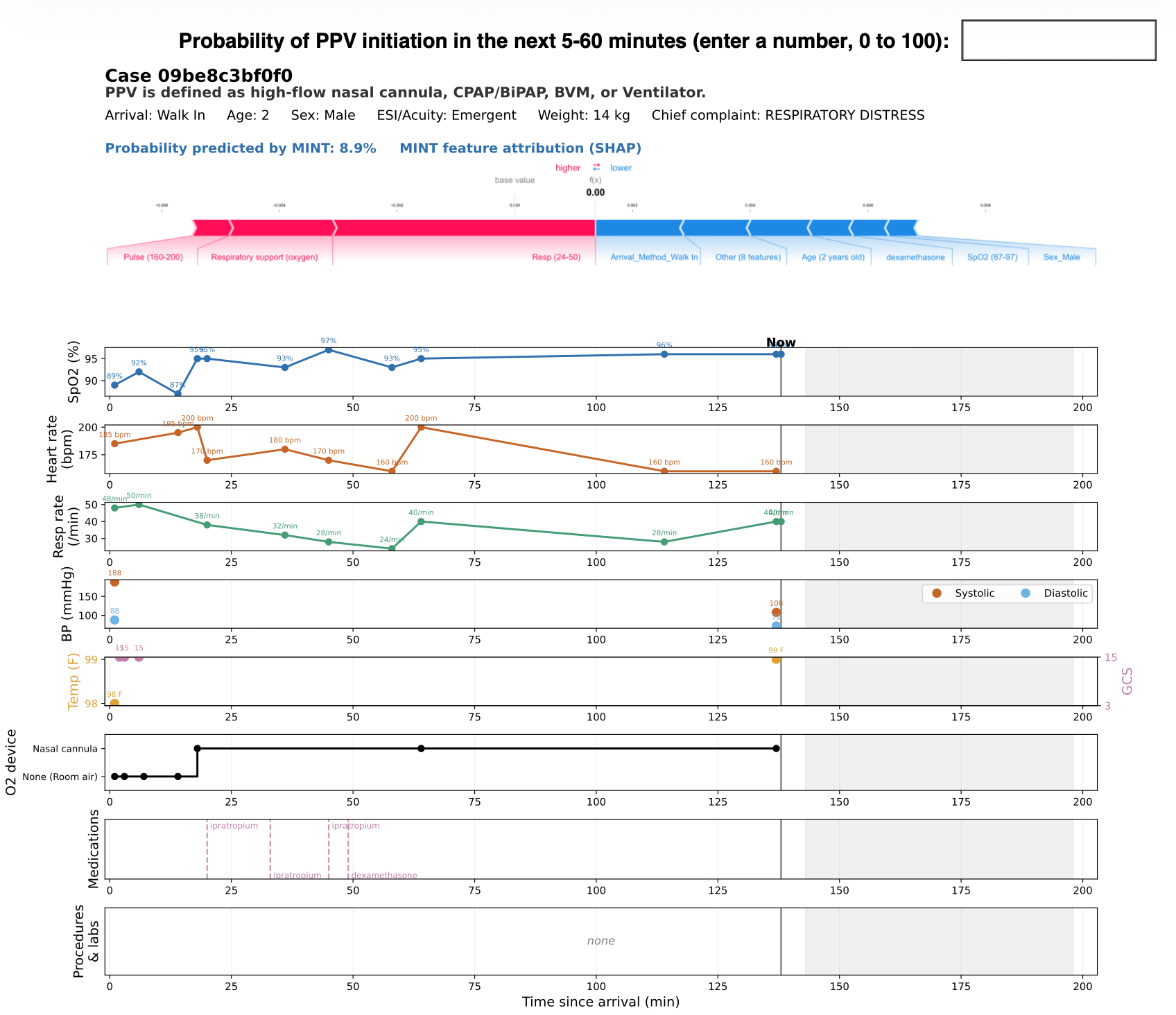
